# Community-Led Diagnosis of Urogenital Schistosomiasis Using a Low-Cost, Point-of-Care Microscopy Toolkit in Rural Nigeria: A mixed-methods study

**DOI:** 10.64898/2026.03.19.26348783

**Authors:** William C. McCarthy, Caroline J. Crain, Tope Olubodun, Ifeoluwa A. George, Simon L. Birk, Uwem F. Ekpo, Hammed Mogaji, Hope T. Leng, Rakesh Kathiresan, Cristina Salas, Morgan H. Sekou, Islamiat Soneye, Makinde Adebayo Adeniyi, Jonathan Beaubrun, Kenechukwu Obumneme Samuel Nwosu, Adedayo Oludolamu, Maryam Kafil-Emiola, Bukunmi Biola Okesola, Phillip J. Koether, Sabona B. Simbassa, Nehali Shah, Megan K. Ngai, Olajide Blessing Oluwanifemi, Iyawe Efosa, Adelawe E. Hassan, Vivian Fagbohun, Boluwatife D. Oladokun, Cutler Cannon, Faneye Oncho, Mahwish Rehman, Avoseh Adeola, Aderionokun J. Stella, Akanbi Abiodun, Kareem Naimot, Sanni Tajudeen Adeola, Olubukola Adelakun, Thomas Copeland, Daniel Amao, Vaibhav Shokeen, Anesta Kothari, Kristina Tebo, Jayhun Lee, Manu Prakash

**Affiliations:** McGovern Medical School, University of Texas Health Science Center at Houston, Houston, TX, USA; Health in Your Hands Diagnostics, Houston, TX, USA; Department of Community Medicine and Primary Care, Federal Medical Center, Abeokuta, Ogun State, Nigeria; Federal University of Agriculture, Abeokuta, Ogun State, Nigeria; Department of Behavioral and Applied Social Sciences, Marian University, Indianapolis, IN, USA; Department of Bioengineering, Stanford University, Stanford, CA, USA; John Sealy School of Medicine, University of Texas Medical Branch, Galveston, TX, USA; Division of Tropical Medicine, Department of Pediatrics, Baylor College of Medicine, Houston, TX, USA; Department of Public Health, Ogun State Ministry of Health, Abeokuta, Ogun State, Nigeria; Department of Systems Engineering, Cornell University, Ithaca, NY, USA; Department of Epidemiology and Human Genetics, University of Texas Health Science Center at Houston School of Public Health, Houston, TX, USA; Department of Microbiology and Molecular Genetics, McGovern Medical School, The University of Texas Health Science Center at Houston, Houston, TX; The University of Texas MD Anderson Cancer Center, UTHealth-Houston Graduate School of Biomedical Sciences; College of Medicine, University of Ibadan, Ibadan, Nigeria; Fountainbridge Humanitarian and Leadership Initiatives, Abeokuta, Ogun State, Nigeria; Jinnah Medical And Dental College, Karachi, Pakistan; Ashoka University, Sonipat, Haryana, India

## Abstract

**Background:** Urogenital schistosomiasis is a major cause of preventable morbidity, primarily in rural, resource-limited regions. After decades of mass drug administration, changing epidemiologic landscapes, and ongoing resource limitations, test-and-treat models may be necessary to meet elimination goals. However, diagnostic capacity remains centralized and laboratory-dependent, and community-led, contextually adapted implementation strategies remain poorly defined. This study describes the accuracy and feasibility of a low-cost diagnostic toolkit and explores community-integrated implementation models.

**Methodology/Principal Findings:** This mixed-methods study enrolled 418 participants from five endemic sites near Oyan River Dam, Ogun State, Nigeria in July 2025. Urine samples underwent parallel analysis by community health extension workers utilizing the toolkit and by laboratory technicians using standard microscopy. The toolkit consisted of a reusable urine filtration device paired with a under-$2 paper microscope. Semi-structured interviews with community health extension workers and key informants were analyzed using the Consolidated Framework for Implementation Research. Prevalence was 27.5% (115/418). Community health extension workers demonstrated progressive improvement in diagnostic accuracy across five sequential communities (n=237), rising from 52.5% (95% CI 37.5–67.1) to 92.1% (79.2–97.3) over eight study days (Cochran-Armitage Z=3.08, p=0.002). Specificity improved from 53.6% to 96.3% (Z=3.00, p=0.003), final sensitivity reached 81.8% (52.3–94.9), and final Cohen’s kappa reached 0.803. In the hands of laboratory scientists, Foldscope microscopy achieved 91.0% sensitivity and 99.3% specificity.

**Conclusions/Significance:** Community-led diagnostic task-shifting for urogenital schistosomiasis control is accurate, feasible, and implementation-ready. Consolidated Framework for Implementation Research-guided analysis demonstrated strong end-user acceptability, with local ownership, collaboration, and trust-building as key implementation facilitators. This approach addresses diagnostic gaps in resource-limited endemic settings with relevance to other community health worker-led strategies.

**Author Summary:** Schistosomiasis is a parasitic infection that spreads through contact with freshwater and often goes undetected and untreated for years. Most common in sub-Saharan Africa, the disease damages the bladder and genitourinary tract, increasing risk of infertility, bladder cancer, and HIV transmission. It is most prevalent in rural communities where the snail intermediate host thrives in local water sources used daily for fishing, farming, and bathing. One such area is the Oyan River in Nigeria. Here, we found that barriers to diagnosis and treatment of the illness include distance and transportation. In this study, community health workers diagnosed their neighbors and community members using a low-cost toolkit: a <$2 / ₦2700 microscope, called the Foldscope paired with a small steel filter card we designed, called the SchistoFilter.. We enrolled 418 people across five villages along the Oyan River in Nigeria and trained eight community health workers to use this toolkit at the point of care. By the fifth community visite, they reached 92.1% accuracy. The study team interviewed community health workers and government officials to contextualize this approach, and they were enthusiastic: The tools can be used with confidence, the training is feasible, and what is most needed is a reliable supply chain and supportive oversight.

## INTRODUCTION

The Oyan River Dam region in Ogun State exemplifies the persistent challenge of urogenital schistosomiasis in Nigeria’s rural communities (1–3). For decades, transmission has been driven by water exposure from vital activities such as fishing, farming, and domestic water use (4,5). Current diagnostic approaches are centralized and laboratory dependent, posing logistical and structural barriers (6–8). Consequently, in the absence of an accessible diagnostic approach to complement mass drug administration (MDA) strategies, many infected individuals remain undiagnosed and untreated, allowing progression to chronic morbidity, including bladder cancer, reproductive tract disease, and increased HIV transmission risk (9–13).

Despite these complex barriers to care, numerous opportunities have emerged to expand access to field-friendly diagnostics. The Foldscope is a commercially available, electricity-free, paper-based microscope whose portability, durability, and ease of use make it particularly suitable for resource-constrained environments (14). The Foldscope has been used for UGS diagnosis, but never by non-microscopists at the point of care (15). To accompany the Foldscope, we developed and, in this study, seek to validate the SchistoFilter, a reusable stainless-steel mesh filter card equipped to trap eggs for visualization under the Foldscope. Together, these components form a reusable diagnostic toolkit that circumvents the resource, cost, and personnel requirements of gold-standard testing, with a total unit cost under $4 USD (approximately ₦5600) and an estimated per-test cost of $0.02-0.04 USD (₦27-56).

Effective implementation of this toolkit requires a community-embedded end-user. Community health extension workers (CHEWs) are trusted and integrated members of rural communities with established roles in public health interventions (16). As community members, CHEWs are equipped to address the numerous structural barriers that currently impact the ability to test and treat UGS (17,18). However, diagnostic testing, particularly those requiring microscopic evaluation, is outside the current scope of CHEW’s daily work. Thus, we sought to evaluate the opinions and perspectives of the CHEWs regarding the proposed diagnostic intervention and to initiate task-shifting of urine sample evaluation to CHEWs. Successful implementation of a community-based diagnostic program requires understanding and addressing the perspectives and concerns of the communities it intends to serve (17, 18, 19).

In this study, we introduce the SchistoFilter, a device that mechanically filters eggs from urine for direct observation under the Foldscope. Here, we tested this Foldscope-SchistoFilter combination (hereafter referred to as ‘the toolkit’) when used in rural communities along the Oyan River by CHEWs, and explored pre-implementation qualitative data regarding feasibility and acceptability.

## METHODS

### STUDY DESIGN

We conducted a mixed-methods cross-sectional study involving three distinct components: (1) evaluation of community sociodemographic characteristics that inform implementation, (2) field-based assessment of diagnostic accuracy comparing the toolkit to reference standard, with parallel evaluation by both trained laboratory scientists and community health extension workers (CHEWs), and (3) qualitative assessment of the feasibility and acceptability of the toolkit among CHEWs and key informants.

The study was conducted in five purposively selected communities surrounding the Oyan River Dam in Abeokuta North (Imala Odo, Abule Tintun, and Abule Sikuru) and Odeda (Ibaro and Apojola) Local Government Areas of Ogun State, Nigeria (Figure 2). Site selection was guided by consultation with local health officials and documented schistosomiasis endemicity, as indicated by historical surveys that showed persistent UGS problems and areas of high and low prevalence (4, 5, 13, 19, 20).

**Fig. 1.**
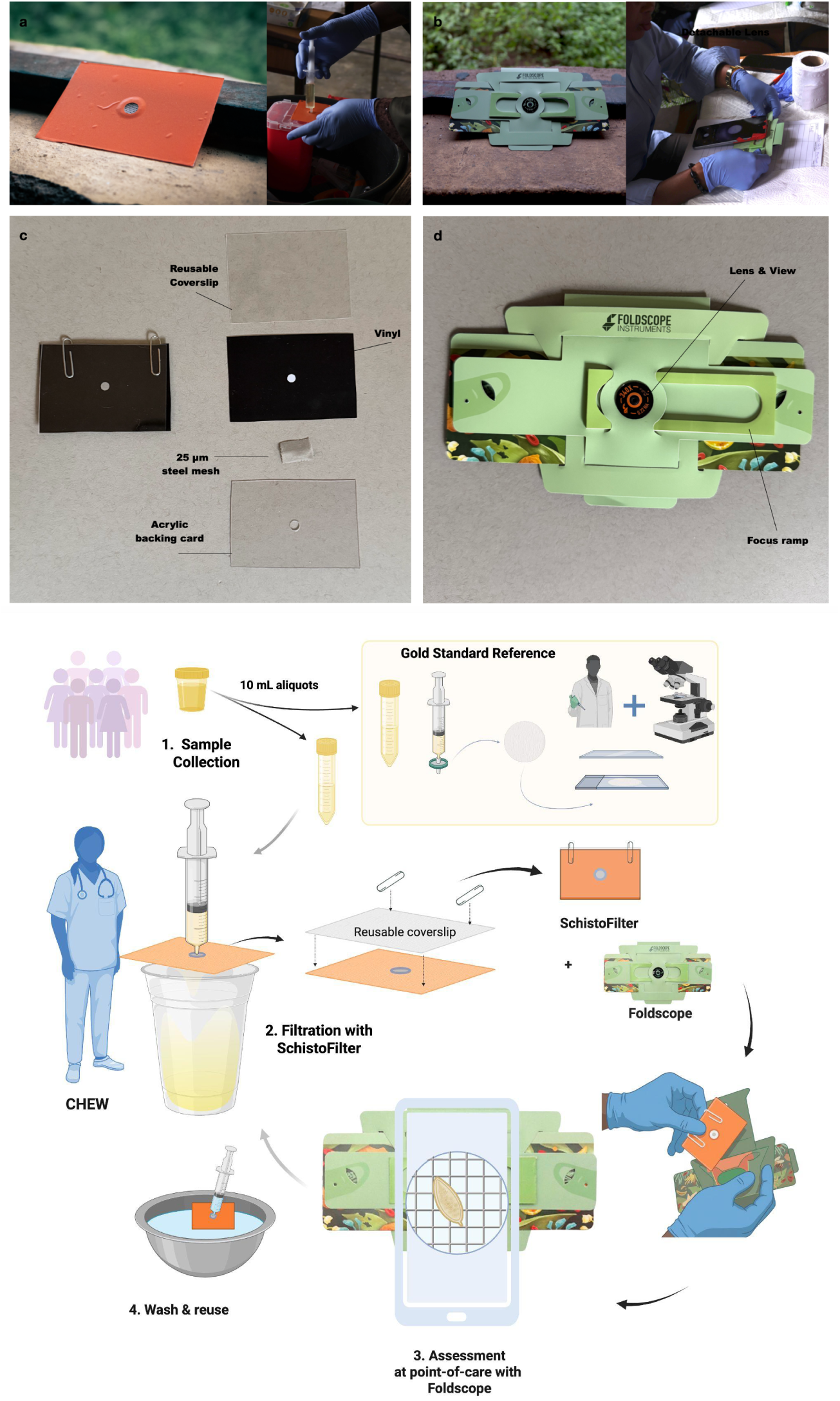
Components of the Foldscope-SchistoFilter toolkit. **(a)** The SchistoFilter is a reusable stainless-steel mesh filter card designed to trap *Schistosoma haematobium* eggs for visualization under the Foldscope. **(b)** The Foldscope is a commercially available paper microscope. When the SchistoFilter is placed inside the Foldscope, schistosome eggs may be visualized directly through the viewing lens or via magnetic smartphone attachment. **(c)** Components of the SchistoFilter prior to assembly. The 25 µm steel mesh was sealed between two acrylic backing cards and covered with orange or black vinyl for durability. A reusable PETG coverslip is secured with paperclips. **(d)** The Foldscope 2.0 with 340× lens, achieving resolution up to 2 microns. Samples are viewed directly or via magnetic smartphone coupling. The optional magnetic, battery-powered LED light module is not shown. See Supplementary Methods for material and construction instructions. **Bottom:** Schematic of parallel sample analysis protocol.

**Fig. 2.**
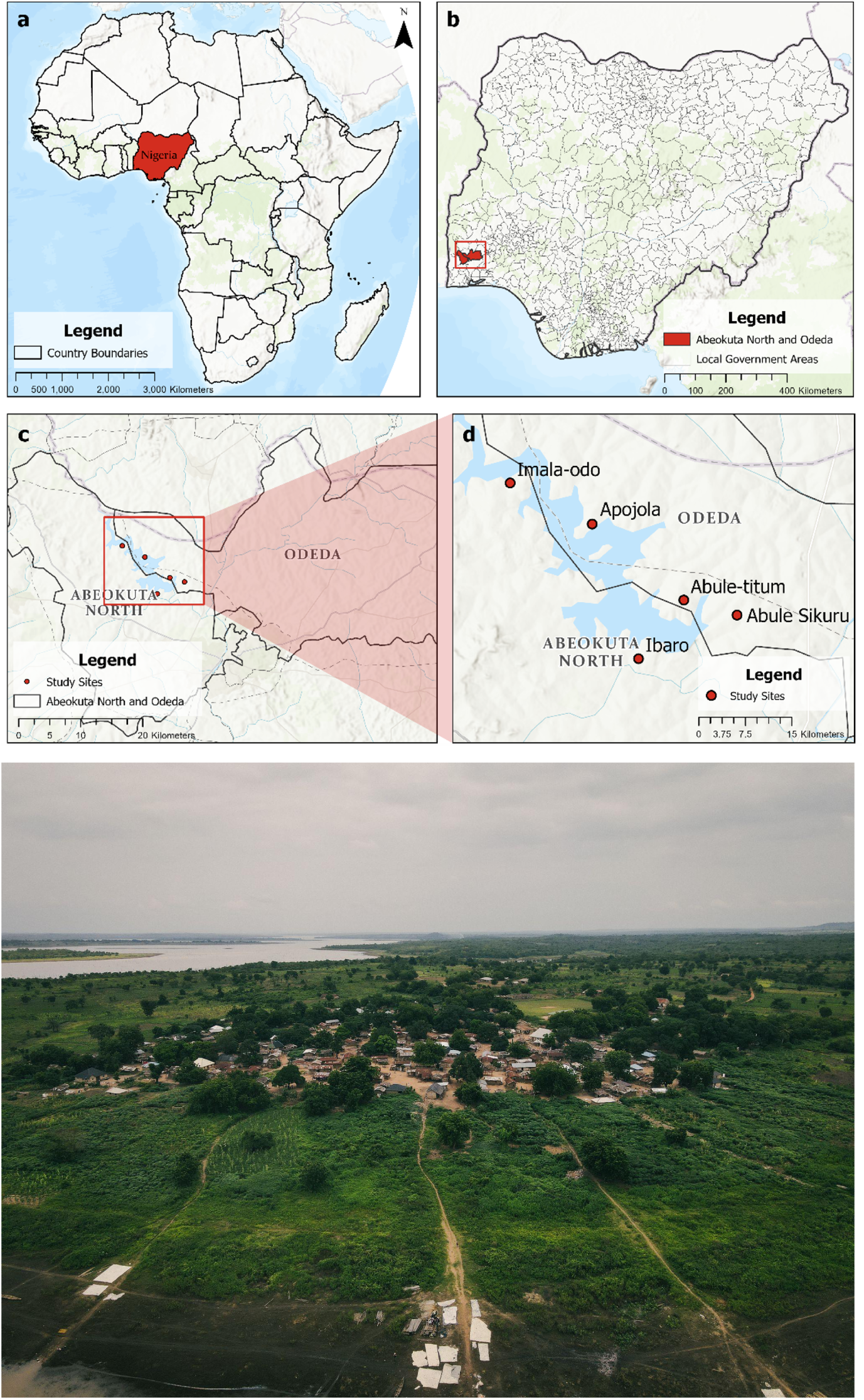
Study area and community locations. Geographic context of the field study conducted in five endemic communities along Oyan River Dam, Ogun State, Nigeria. **(a)** Reference map showing Nigeria’s location in West Africa. **(b)** Ogun State highlighting Abeokuta North and Odeda Local Government Areas (LGAs) where the study was conducted. **(c)** Detailed view of study site locations within the LGAs, showing proximity to Oyan River Dam. **(d)** Individual study sites: Imala Odo, Ibaro, Apojola, Abule Titun, and Abule Sikuru. All communities are situated along waterways where daily water contact for domestic, agricultural, and fishing activities sustains schistosomiasis transmission. **Bottom:**Drone aerial photograph of Imala Odo village illustrating the remote, rural character of study communities. Mapping conducted using ArcGIS Pro 3.3.0 (Esri, Redlands, CA) with administrative boundaries from GRID3 (grid3.org).

Ethical approval was granted prior to study initiation by the Ogun State Health Research Ethics Committee (OGHREC/467/2025/618/APP) and The University of Texas Health Science Center at Houston (HSC-MS-25-0502). Community sensitization visits were conducted with support from the Ogun State Neglected Tropical Disease (NTD) Office, and verbal approval was provided by community leaders. Each participant gave informed consent with a physical or electronic signature.

### PARTICIPANTS

In each community, residents aged five and older were recruited at a preferred location within the community as suggested by local leaders. The target sample size was 365 participants, calculated to estimate the sensitivity and specificity within a 5-percentage point margin of error at 95% confidence (See Supplementary Materials for detailed calculation). 418 community members were ultimately recruited.

Following documented consent, recruited community members were asked to provide at least 30 mL of urine for analysis using (1) a reference standard and (2) the Foldscope-SchistoFilter toolkit (see Figure 3). At the time of sample collection, participants were invited to complete a demographic questionnaire to assess socioeconomic factors impacting schistosomiasis endemicity, diagnosis, and treatment, as well as their individual knowledge of schistosomiasis. The questionnaire was administered verbally in participants’ preferred language by trained research team members. Responses were documented on paper (n=50) or online via SurveyCTO (n=368).

**Fig. 3.**
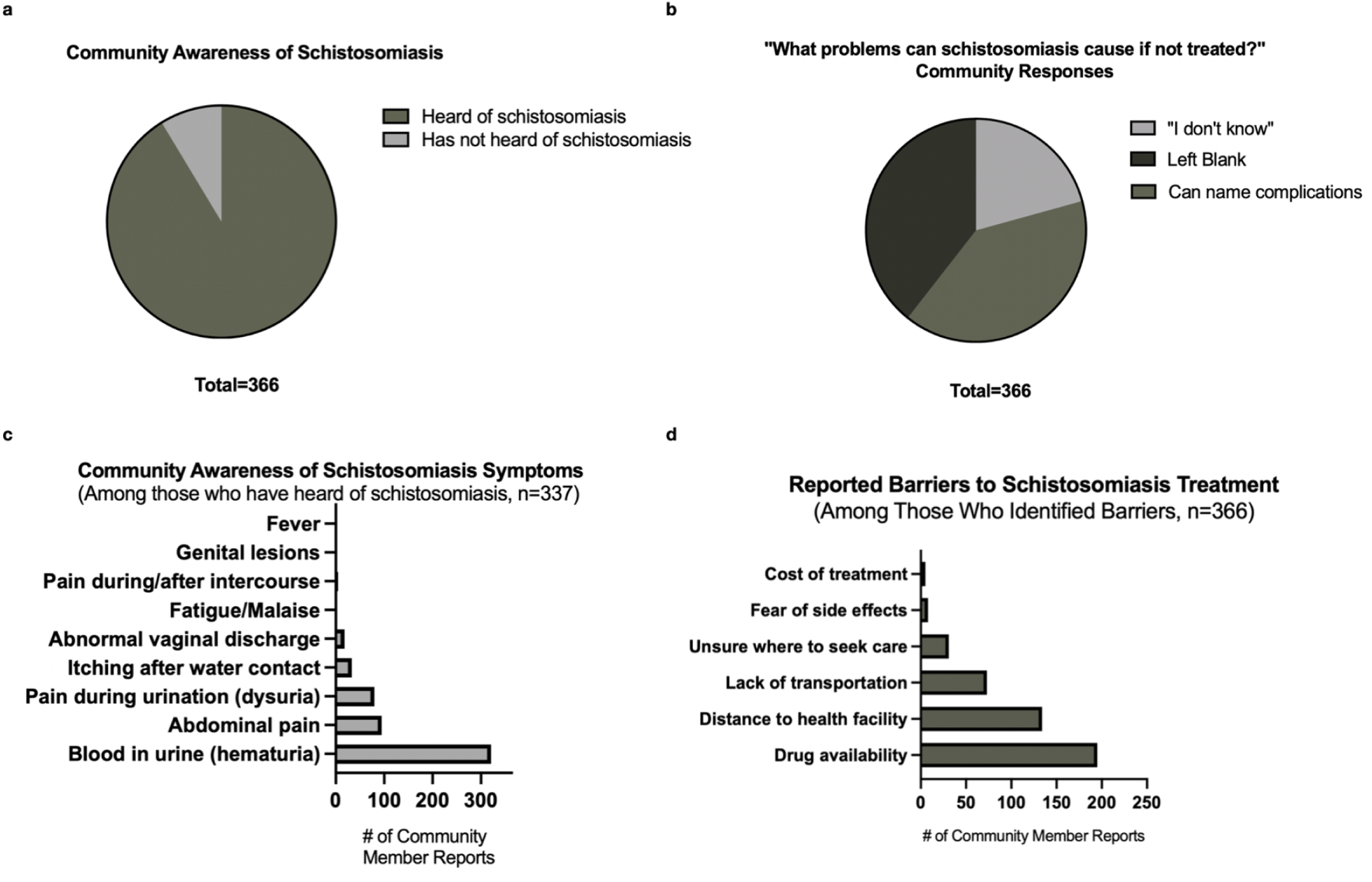
Community knowledge of schistosomiasis and barriers to treatment. Assessment of disease awareness, symptom recognition, complication knowledge, and treatment barriers among 366 participants with complete demographic data from five communities along Oyan River Dam. **(a)** Disease awareness: 334/366 (91.3%) had heard of schistosomiasis or its Yoruba name ‘Atosi Aja’ (bloody urine). **(b)** When asked to name complications of untreated schistosomiasis, only 146/366 (39.9%) provided substantive answers, while 76/366 (20.8%) stated ‘don’t know’ and 144/366 (39.3%) left the question blank, revealing substantial gaps in complication knowledge despite high disease awareness. **(c)** Symptom recognition among those aware of schistosomiasis (n=334): hematuria was recognized by 96.1% of aware respondents; recognition of other symptoms was substantially lower: blood in stool (53.9%), abdominal pain (28.4%), dysuria (24.0%), itching after water contact (9.9%), and genital symptoms (7.5%). **(d)** Barriers to treatment access among 366 participants: drug availability (53.3%), distance to health facility (36.6%), and lack of transportation (19.9%) were most frequently cited; knowledge gaps and cost were cited less frequently.

A total of 418 community members were enrolled and provided urine samples for diagnostic testing (Supplemental STROBE Diagram). Of these, 366 (87·6%) completed entire demographic questionnaires; 52 participants (12·4%) had incomplete demographic data due to problems syncing digital forms or incomplete responses. For the CHEW field validation component, 237 participants (56·7% of enrolled) were used for the diagnostic accuracy analysis. The remaining 181 samples were excluded from CHEW validation analysis due to time constraints and field practicalities of a reasonable workload for the CHEWs. Following the field study, trained laboratory scientists blinded to results examined 412 intact glass slides (98·6% of 418 collected; 6 excluded due to damage during transport) using Foldscope microscopy, evaluating the same membrane filters previously analyzed by gold-standard compound microscopy.

CHEWs were recruited from study sites and surrounding communities. All CHEWs primarily served rural communities and had no prior microscopy experience.

Information on additional participants, CHEWs recruitment and participation, and participant compensation can be found in the Supplemental Appendix.

### PROCEDURES

Over a period of eight study days (See timeline in Supplemental Appendix, Supplemental Figure 1), CHEWs received 2-3 hour training sessions on the first three days, focusing on sample preparation and egg identification by size, shape, and color.

Trained laboratory technicians conducted the reference standards. Following filtration of the 10 mL aliquot using a 20 µm polycarbonate membrane filter (Sterlitech) secured within a filter holder (Sterlitech). After filtration, the membrane was placed on a glass slide and examined under a compound light microscope (AmScope B120 Compound Microscope 10-40X objectives).

Participating CHEWs conducted the experimental arm. A 10-mL aliquot was drawn from the same urine collection cup into a Luer-lock disposable syringe (BH Supplies BH110L) and slowly expelled through the central mesh aperture of the SchistoFilter (Figure 1A) into a small biohazard container. The SchistoFilter was then inserted into the Foldscope and examined using an affordable mobile phone, TecnoPop 10 (Tecno Mobile), with a 13-megapixel camera, the minimum recommended resolution for use with Foldscope (21). The phone camera is paired to the Foldscope via magnetic attachment as part of the Foldscope 2.0 kit, reinforced with tape if needed. After visualization, the SchistoFilter was washed in two sequential bowls containing packaged sterile sachet water, dried on clean paper towels, and prepared for reuse.

Detailed explanations of the filtration and microscopy procedures, as well as subsequent statistical analyses, can be found in the Supplemental Materials.

### SECONDARY ANALYSES

After the completion of the field study, reference standard slides containing Sterlitech-filtered patient samples were formalin-preserved and analyzed by cascade-trained laboratory scientists using the Foldscope. Diagnostic accuracy metrics, including time-to-assessment and egg count, were reported

### QUALITATIVE DATA COLLECTION AND ANALYSIS

This study was designed to assess the technical performance of the toolkit and evaluate its fit within the social, institutional, and material realities of the setting in which it would be used. Accordingly, we integrated qualitative and implementation science methodologies from the outset to examine how local workflows, resource constraints, power dynamics, and user experiences shape the future feasibility and acceptability of our toolkit.

Among the CHEWs who participated in the quantitative validation of the toolkit, four were available to participate in semi-structured interviews following the conclusion of the field study. Furthermore, four key informants, including local and regional NTD leadership, were recruited to participate in semi-structured interviews.

Coding of verbatim-transcribed transcripts was conducted using a combined deductive and inductive framework based on Consolidated Framework for Implementation Research (CFIR) constructs and emergent themes, respectively. Additional constructs related to implementation power dynamics (22) were included in order to strengthen the framework’s relevance and analytical depth. Salient constructs were identified based on frequency of code application, relevance, and richness of data.

### DATA MANAGEMENT

Complete demographic data was available for 366/418 (87.6%) participants enrolled. Missing demographic data arose from software-related technical difficulties for 52 participants. These data losses primarily occurred during days one and two of data collection but were otherwise unrelated to participant characteristics or diagnostic outcomes. Please refer to the “DATA MANAGEMENT” Section of the Supplemental Appendix for more information on data organization.

### STATISTICAL ANALYSIS

Quantitative data were analyzed using GraphPad Prism version 10.5 (GraphPad Software, San Diego, CA, USA) and R version 4.5.2 (R Foundation for Statistical Computing, Vienna, Austria) via Posit Cloud (Posit PBC, Boston, MA, USA). Statistical significance was set at p<0.05 unless otherwise specified.

Overall prevalence and prevalence stratified by demographic characteristics were calculated as proportions with 95% confidence intervals (CIs) using the Wilson score method. Chi-square tests evaluated associations between categorical variables (sex, occupation, community) and infection status. One-way ANOVA compared continuous variables (age, egg counts) across multiple groups, with post-hoc pairwise comparisons using Tukey’s honestly significant difference test. Pearson correlation coefficient (r) assessed the relationship between age and infection intensity (egg counts per 10 mL urine) after confirming approximate normality of residuals.

Sensitivity, specificity, positive predictive value (PPV), negative predictive value (NPV), and overall accuracy were calculated from standard 2×2 contingency tables, with the WHO-approved Sterlitech membrane filtration microscopy as the reference standard. Wilson score 95% CIs were used for all proportions. Cohen’s kappa (κ) assessed agreement between CHEW assessments and the reference standard (κ<0.20, poor; 0.21–0.40, fair; 0.41–0.60, moderate; 0.61–0.80, substantial; 0.81–1.00, almost perfect). Fisher’s exact test (two-tailed) evaluated the statistical significance of association between index test and reference standard. Positive and negative likelihood ratios were calculated to assess clinical utility (LR+ >10 and LR− <0.1 indicating strong diagnostic value).

### ROLE OF THE FUNDING SOURCE

Study sponsors had no role in study design, data collection, analysis, interpretation, or the decision to submit for publication.

## RESULTS

### DEMOGRAPHIC CHARACTERISTICS OF STUDY PARTICIPANTS

This study enrolled 418 participants from five communities along the Oyan River Dam. Complete demographic data were collected for 366 participants (87.6% of the total participants recruited), meeting the target sample size (Table 1). Of these 366 participants, 53.1% were female, and sex distribution did not differ significantly across study sites (p=0.12).

**Table 1.** Community Member Demographics Survey Results. N = 366. Complete demographic data were collected for 366 participants (87.6% of 418 enrolled), meeting the prospectively calculated target sample size of 365. Data are n (%). Sex distribution did not differ significantly across study sites (p=0.08). Incomplete data were excluded from analysis.

| Characteristic | n (%) |
| --- | --- |
| **Age (years)** |  |
| 5–15 | 146 (39.9) |
| 16–25 | 42 (11.5) |
| 26–45 | 108 (29.5) |
| 46–93 | 70 (19.1) |
| **Sex** |  |
| Male | 174 (47.5) |
| Female | 192 (52.5) |
| **Occupation** |  |
| Student (unemployed) | 163 (44.5) |
| Farming | 84 (23.0) |
| Fishing | 64 (17.5) |
| Artisan | 21 (5.7) |
| Other | 28 (7.7) |
| Civil servant | 4 (1.1) |
| Housewife | 2 (0.5) |
| **Community** |  |
| Imala Odo | 123 (33.6) |
| Ibaro | 63 (17.2) |
| Apojola | 104 (28.4) |
| Abule Titun | 51 (13.9) |
| Abule Sikuru | 25 (6.8) |

Barriers to treatment access were identified through the questionnaire. Among 174 reported barriers to treatment access, knowledge gaps (29.9%) and drug availability (27.0%) emerged as the most frequently cited obstacles, followed by distance to health facilities (16.1%), highlighting critical gaps in treatment accessibility in these endemic communities (Table 1).

### AWARENESS OF SCHISTOSOMIASIS & BARRIERS TO TREATMENT

A total of 85.1% of participants were able to recognize schistosomiasis or *Atosi Aja*, Yoruba for “Bloody urine” (Fig. 3A). When asked to identify problems caused by untreated schistosomiasis, study participants’ responses (N=366) fell into three categories: 146/366 (39.9%) provided substantive answers such as “infertility”, “death”, “stunted growth”, or “blood loss”; 76/366 (20.8%) explicitly stated, ‘I don’t know’; and 144/366 (39.3%) left the question blank. Awareness of symptoms was strong for hematuria, lacking for other key symptoms (Figure 3C).

A total 92.4% of participants reported at least one obstacle to care, with drug availability (53.3%), distance to health facility (36.6%), and lack of transportation (19.9%) being the most commonly cited obstacles (Figure 3C). The remote, rural characteristics of the study communities is represented in the drone aerial image (Figure 2).

### PREVALENCE OF SCHISTOSOMIASIS ACROSS STUDY COMMUNITIES

Of the 418 participants tested using gold-standard membrane filtration microscopy, 115 (27.5%) tested positive for *Schistosoma haematobium* infection. To collect this prevalence data, the WHO-approved gold standard of a Sterlitech filter and a syringe system for filtering urine was used. The Sterlitech filter system costs $1.50 (2170 NGN) per test, excluding urine collection cups.

Participants aged 15-24 years showed the highest infection rates (53.7%). Infection intensity (Fig. 4B), measured by egg counts per 10 mL urine, showed a significant but weak negative correlation with age (Pearson’s r = -0.17, p=0.0013), with most affected adults over 40 years showing light infections (<10 eggs/10 mL). Infection rates varied by occupation, with students representing the largest burden of positive cases (Supplementary Figure 2). A full description of prevalence by age and occupation is presented in Supplementary Table 3.

**Fig. 4.**
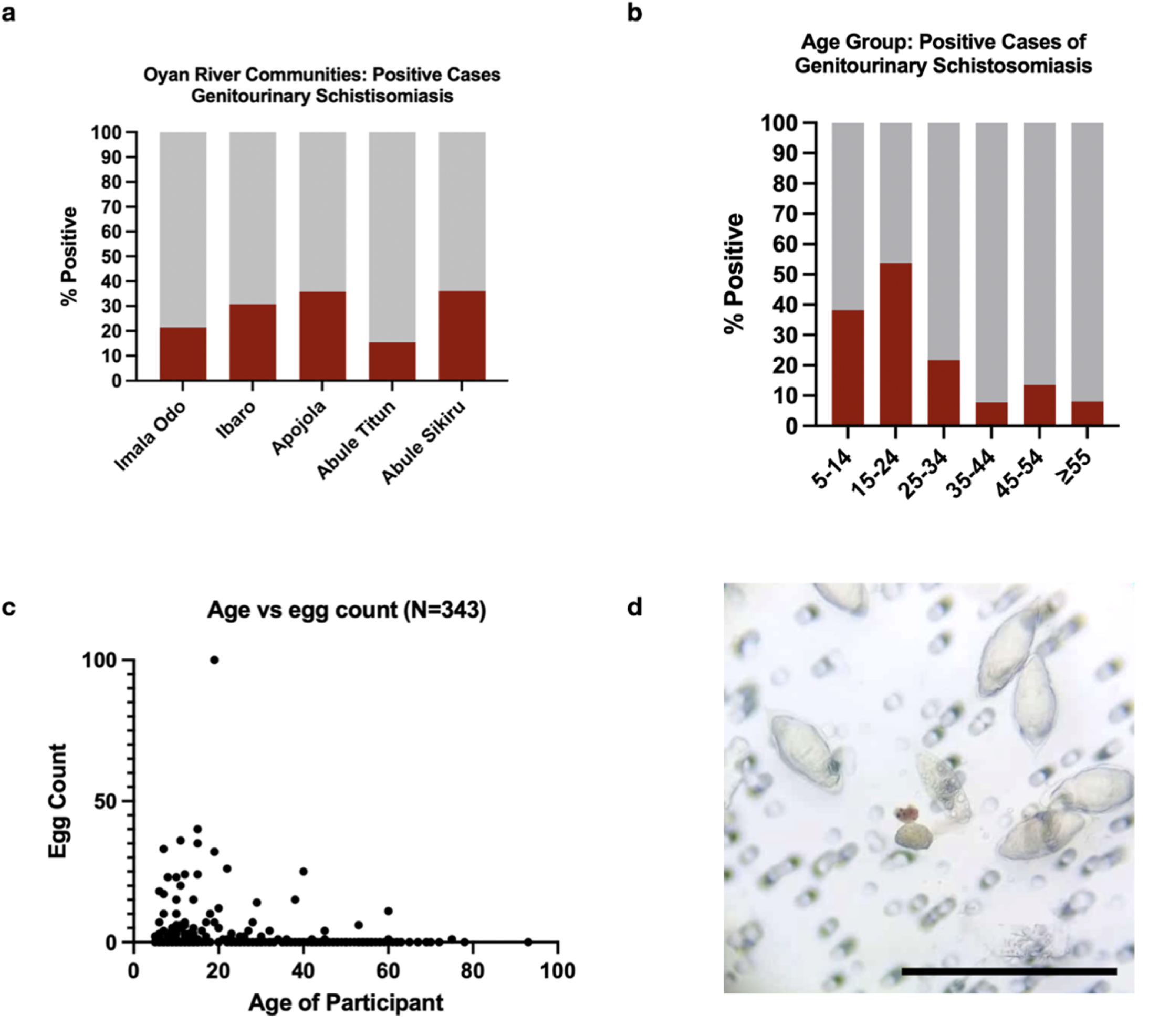
Prevalence and age distribution of urogenital schistosomiasis. Epidemiological findings from reference standard microscopic examination of 418 urine samples across five study communities. **(a)** Community prevalence by gold-standard membrane filtration microscopy: Apojola (35.8%, n=106), Abule Sikuru (36.0%, n=50), Ibaro (30.8%, n=65), Imala Odo (21.4%, n=145), and Abule Tuntun (15.4%, n=52). Overall prevalence 115/418 (27.5%). Prevalence differed significantly across communities (χ²=12.42, df=4, p=0.015). **(b)** Age-specific prevalence: highest infection rates in 15–24 year olds (53.7%), followed by 5–14 years (35.6%), 25–44 years (28.7%), and ≥45 years (14.3%). **(c)** Infection intensity by age: egg counts per 10 mL urine plotted against participant age (n=418). Highest egg burdens in children and adolescents (up to 100 eggs/10 mL in the 0–20 age range) with progressive decline in adults. Significant but weak negative correlation between age and infection intensity (Spearman r=−0.17, p=0.0013). Most infected adults >40 years showed light infections (<10 eggs/10 mL). **(d)** Representative microscopy image showing characteristic *Schistosoma haematobium* eggs (terminal spine visible) identified in participant urine samples by gold-standard light microscopy. Scale bar, 400 µm.

### FIELD VALIDATION OF POINT-OF-CARE DIAGNOSTIC KIT PERFORMED BY CHEWS

A total of 237 randomly selected urine samples from participants across five sequential communities (eight total days) underwent parallel testing: gold-standard membrane filtration microscopy performed by trained laboratory scientists and point-of-care assessment using the toolkit operated by CHEWs in the field (Table 2,3).

**Table 2.** CHEW Diagnostic Performance Across Sequential Communities Using the Foldscope-SchistoFilter Toolkit. N = 237. Cochran-Armitage trend test demonstrated significant improvement for accuracy (Z=3.08, p=0.002) and specificity (Z=3.00, p=0.003). Sensitivity showed a positive but non-significant trend (Z=1.04, p=0.301). Overall pooled performance: sensitivity 62.7% (95% CI: 51.4–72.7%), specificity 74.7% (67.5–80.8%), accuracy 70.9% (64.8–76.3%). Wilson score 95% confidence intervals. Full metrics in Supplementary Table S3.

| Sequentially Visited Village (L to R) | Imala Odo | Ibaro | Apojola | Abule Tintun | Abule Sikuru |
| --- | --- | --- | --- | --- | --- |
| Sample Size | 40 | 42 | 87 | 30 | 38 |
| Accuracy, % (95% CI) | 52.5 (37.5–67.1) | 76.2 (61.5–86.5) | 69.0 (58.6–77.7) | 66.7 (48.8–80.8) | 92.1 (79.2–97.3) |
| Sensitivity, % (95% CI) | 50.0 (25.4–74.6) | 78.6 (52.4–92.4) | 53.1 (36.4–69.1) | 66.7 (30.0–90.3) | 81.8 (52.3–94.9) |
| Specificity, % (95% CI) | 53.6 (35.8–70.5) | 75.0 (56.6–87.3) | 78.2 (65.6–87.1) | 66.7 (46.7–82.0) | 96.3 (81.7–99.3) |
| Cohen's kappa | 0.031 | 0.500 | 0.319 | 0.242 | 0.803 |

**Table 3.** Performance of the Toolkit Relative to the Reference Standards. N = 237. Communities are listed in sequential order in which they were visited. Thus, community number five (Abule Sikuru) represents the day at which community health workers had maximum training and experience. The Complete diagnostic metrics: PPV, NPV, Cohen’s kappa, likelihood ratios, and operational measures are presented in Supplementary Table S3.

|  | Reference Standard (+) | Reference Standard (-) | **Total** |
| --- | --- | --- | --- |
| Foldscope-SchistoFilter (+) | 47 | 41 | 88 |
| Foldscope-SchistoFilter (-) | 28 | 121 | 149 |
| **Total** | 75 | 162 | 237 |

Community health extension workers demonstrated progressive improvement in diagnostic agreement with the gold standard across sequential deployments (Table 2). Accuracy improved from 52.5% (95% CI: 37.5-67.1) at the first community (Imala Odo, n=40) to 92.1% (95% CI: 79.2-97.3) at the fifth community (Abule Sikuru, n=38), demonstrating statistically significant linear improvement (Cochran-Armitage trend test: Z=3.10, p=0.002). Cohen’s kappa improved from 0.031 (poor agreement) at Imala Odo to 0.803 (substantial agreement) at Abule Sikuru, indicating progression from essentially random performance to expert-level diagnostic capability (19). Error rates declined significantly across the study period, with a statistically significant negative correlation between sample number and error rate (Fig. 5, Spearman ρ = -0.830, p<0.001).

**Fig. 5.**
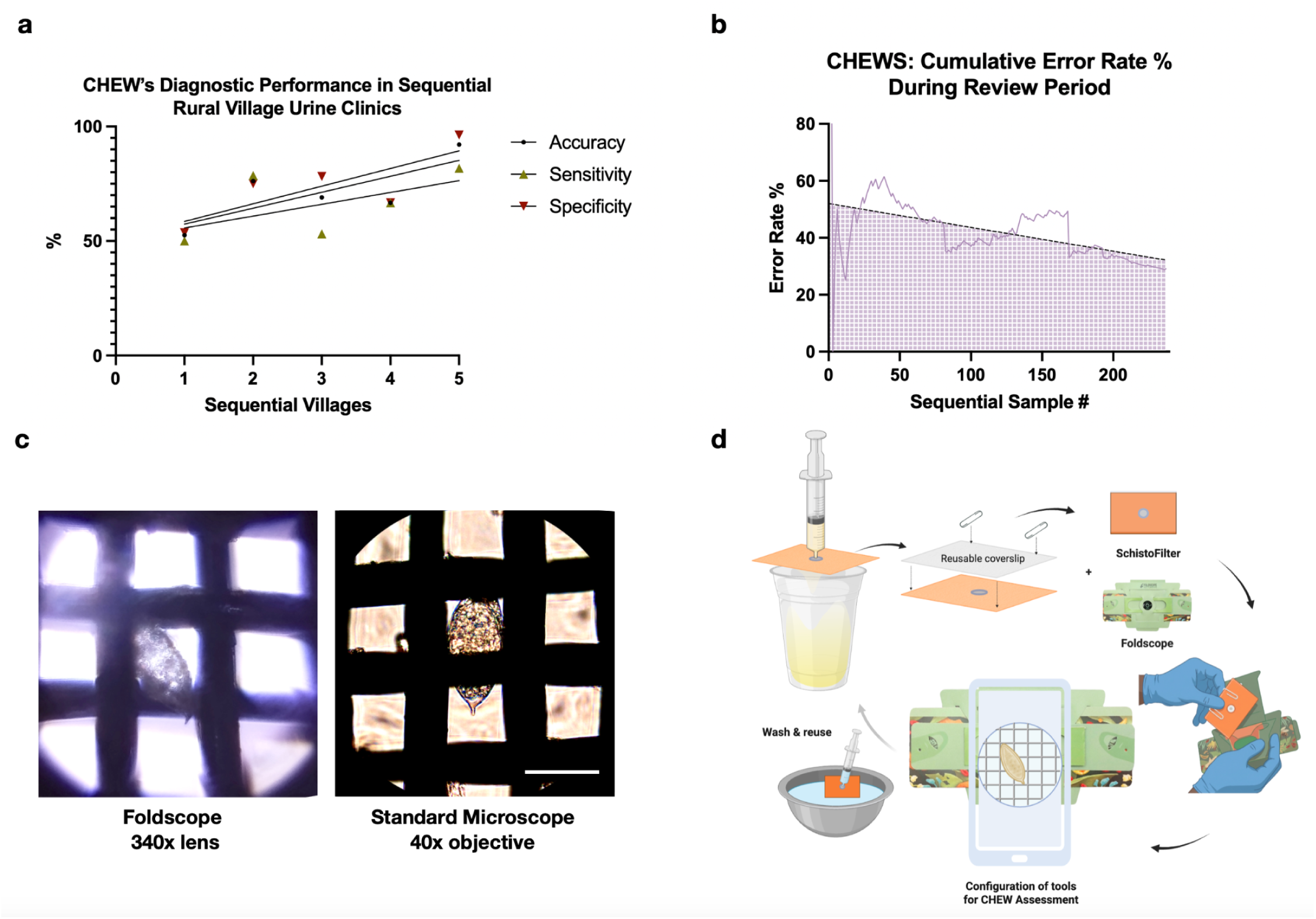
Progressive improvement in CHEW diagnostic performance. Learning curve demonstrating systematic improvement in community health extension worker (CHEW) diagnostic accuracy across five sequential community screenings (n=237 samples, 8 study days). **(a)** Diagnostic performance metrics by site: sensitivity (open green triangles) improved from 50.0% to 81.8%, specificity (open red inverted triangles) from 53.6% to 96.3%, and overall accuracy (filled black circles) from 52.5% to 92.1%. Solid lines represent linear regression fits (sensitivity: R²=0.32; specificity: R²=0.60; accuracy: R²=0.58). Cochran-Armitage trend tests demonstrated statistically significant improvement for accuracy (Z=3.08, p=0.002) and specificity (Z=3.00, p=0.003); sensitivity improvement was non-significant (Z=1.04, p=0.301). **(b)** Cumulative error rate by sequential sample number (n=237). Error rates declined significantly throughout the study period (Spearman ρ=−0.830, p<0.001), stabilizing below 10% by sample 200. **(c)** Representative images of *Schistosoma haematobium* eggs filtered with the SchistoFilter and visualized using (left) Tecno Pop 10 smartphone attached to Foldscope 340×lens, compared to (right) standard compound light microscopy. Scale bar, 70 µm.Image quality from smartphone-coupled Foldscope was sufficient for diagnostic decision-making in field conditions. **(d)** Schematic of CHEW sample preparation workflow: (1) urine collection; (2) SchistoFilter filtration; (3)Foldscope examination and smartphone documentation; (4) filter cleaning for reuse. Sample sizes by site: Imala Odo (n=40), Ibaro (n=42), Apojola (n=87), Abule Titun (n=30), Abule Sikuru (n=38).

The average diagnostic turnaround time was approximately 11 minutes per sample (Supplemental Table 1). The field photographs (Supplemental Figure 3) illustrate the point-of-care workflow, with CHEWs conducting sample processing and microscopic examination in the communities’ schoolhouses, demonstrating the feasibility of decentralized diagnostic services in resource-limited environments. Across all 237 assessments, CHEWs demonstrated pooled sensitivity of 62.7% (95% CI 51.4–72.7%), specificity of 74.7% (67.5–80.8%), and overall accuracy of 70.9% (64.8–76.3%). Fisher’s exact test confirmed a significant association between CHEW assessments and reference standard microscopy (p<0.001), with fair overall agreement (Cohen’s κ=0.357).

### SECONDARY ANALYSIS OF FOLDSCOPE PERFORMANCE IN OPTIMAL CONDITIONS

Following the field study, trained laboratory scientists blinded to results examined 412 intact glass slides using Foldscope microscopy, evaluating the same membrane filters previously analyzed by gold-standard compound microscopy. Laboratory-based Foldscope examination demonstrated excellent diagnostic performance: sensitivity 90.99% (95% CI: 84.2-95.0), specificity 99.34% (95% CI: 97.6-99.9), positive predictive value 98.06% (95% CI: 93.2-99.7), and negative predictive value 96.76% (95% CI: 94.2-98.2) (Table 4,5). Fisher’s exact test demonstrated significant agreement between methods (p<0.0001, two-sided). Agreement between Foldscope and conventional microscopy was excellent, with Cohen’s kappa coefficient of 0.918 (32).

**Table 4.** Performance of the Foldscope with SterliTech Filtration Relative to Reference Standard by Laboratory Technicians. N = 412. Complete diagnostic metrics, including PPV, NPV, Cohen’s kappa, likelihood ratios, and operational measures in Supplementary Tables S4,S5.

| **Metric** | **Value** | **95% CI** |
| --- | --- | --- |
| Sensitivity | 90.99% | 84.21–95.03 |
| Specificity | 99.34% | 97.61–99.88 |
| Positive Predictive Value (PPV) | 98.06% | 93.19–99.65 |
| Negative Predictive Value (NPV) | 96.76% | 94.15–98.23 |
| Accuracy | 97.10% | 95.0–98.4 |
| Cohen's Kappa | 0.918 | 0.87–0.96 |

**Table 5.** Performance of the Foldscope with SterliTech Filtration Relative to Reference Standard by Laboratory Technicians. N = 412. Confusion matrix.

|  | Reference Standard (+) | Reference Standard (-) | **Total** |
| --- | --- | --- | --- |
| Foldscope (+) | 101 | 2 | 103 |
| Foldscope (-) | 10 | 299 | 309 |
| **Total** | 111 | 301 | 412 |

Analysis of accuracy throughout sequential samples revealed rapid skill acquisition, with cumulative accuracy stabilizing above 95% after approximately 100 samples and reaching a plateau of 97-98% by sample 200. The time-to-assessment per sample showed high initial variability (range: 5-20 minutes) that progressively decreased and stabilized to 3-8 minutes after 100 samples, with median time declining from approximately 10 minutes to 5 minutes as examiners gained proficiency (Supplemental Fig. 5, p<0.0001, linear regression). The positive likelihood ratio of 137.0 far exceeds the threshold of 10 considered clinically useful, indicating that positive results approached diagnostic certainty.

### QUALITATIVE ASSESSMENT OF FEASIBILITY AND ACCEPTABILITY

#### INNOVATION: Relative Advantage, Innovation Complexity, and Innovation Design

Across the interviews, the toolkit was viewed positively, with its relative advantage emerging as a primary facilitator. CHEWs emphasized that they currently lack access to any diagnostic tools for schistosomiasis, resulting in reliance on empirical treatment or patient referral. In this context, the toolkit was perceived as addressing a critical gap in care by enabling community-based diagnosis, thereby strengthening the reach and legitimacy of patient care.

> “Previously we don’t have anything to diagnose…It would make my work easier to diagnose the patient with schistosomiasis. When a patient complains of passing blood in the urine, after counseling I would be able to tell them that they will be tested.” (CHEW 2)

> “In the community, there is no diagnostic center. So if you suspect, you will treat…The closest diagnostic center is far. It is more than 10 kilometers. It is up to 1.5 hours by car. By bike it would be two hours..It is too far for them to go there. It is too far and it will cost them. Transport, the cost to get themselves down is too much…Some of the people don’t go even if we send them.” (CHEW 3)

Though stakeholders were unanimously enthusiastic about the availability of a diagnostic tool, the complexity of the toolkit emerged as both a barrier and a facilitator.

> “[At the beginning] everything seems very difficult. But after being familiar with it, we see it is very easy to examine and use. Compare what you were doing on the first or second day to what we’re doing today. It is very, very different. We are more comfortable with the use and everything, so it is very easy now.” (CHEW 1)

CHEWs unanimously agreed that they felt confident using the toolkit to screen patients in their communities by the end of the study, further substantiated by the linear increase in accuracy demonstrated in Figure 5. Thus, the gap between anticipated and experienced complexity, particularly in end-users without microscopy experience, highlights the potential importance of early socialization by trusted collaborators to build confidence and self-efficacy.

Finally, while the toolkit was designed to be waterproof, fully reusable, and suitable for field conditions, a prominent concern from CHEWs was its durability. CHEWs expressed hesitancy over the reusability of both the Foldscope and the SchistoFilter, stating:

> “[The concern] is only if we will be using and reusing and reusing and eventually if any of the appliance gets spoiled, we don’t see it very clearly.” (KII 3)

These narratives suggest that early implementation efforts should prioritize training and opportunities for hands-on practice to mitigate usability concerns and reinforce trust in design quality, and that the kit is well-packaged for travel and reuse. The design may also be iterated upon to ensure visual guides and references are incorporated to allow for easy reference to reuse protocols for washing, guidance on the number of uses, and storage.

#### OUTER SETTING: Local Conditions, Partnerships, and Connections

CHEWs’ concerns about durability reflected the practical constraints of their local context. The primary implementation barrier identified in the Outer Setting was related to supply chain sustainability and reliable distribution of consumables. Stakeholders emphasised the difficult nature of restocking, underscoring the importance of fully reusable tools. Juxtaposing the toolkit’s perceived advantages, stakeholders cite sustainability as a particular threat to community and stakeholder trust.

> “Once people have been assessing the use of the Foldscope and they have been seeing the efficacy and suddenly a break in the supply chain arises, people might not have all that much confidence in whatever the health workers are doing again. We want continuity of government to support the use and supply of the Foldscope.” (KII 3)

Key informant interviews indicated that local and state Ministries of Health were capable of managing supply chains and distribution. However, concerns about the potential long-term accessibility of the toolkit by CHEWs underscore the need to strengthen end-user trust in systems built to support implementation.

> “In the NTD control program, we have a logistic unit. So I believe we have all the procurement like we have for other programs in the unit…The logistical [head] of the unit will be notified of the [need] of the procurement of the materials…So after we have gotten all those, we distribute the materials in the local government area (LGA) and to the coordinators.” (KII 1)

#### INNER SETTING: Tension for Change, Mission Alignment, Compatibility, and Discursive Power

A strong tension for change was evident in the CHEW and KII narratives. This tension was primarily driven by interruptions in MDA, leading to growing recognition that reliance on externally donated pharmaceuticals undermines the predictability, autonomy, and efficacy of schistosomiasis control efforts at the local level.

> “But unfortunately, about a year or two ago now, we have not been able to come up with the treatment…We cannot say specifically [why], it just stopped. I don’t know if it was logistic. Other than that, there is none.” (KII 2)

> “In past years, there are no LGAs that we have that we’ve treated. The last time we treated adults and children was in 2023. Also, there is a paucity of medicines; they are donated. There are no medicines. I think it’s [pharmaceutical company name]. [The pharmaceutical company] donates them to the families from the government. So that is something that is also a problem.” (KII 4)

There is a clear frustration with the gaps in drug availability in recent years, with a possible underlying tension of a perceived lack of agency among the Ministry of Health and local NTD teams. This dependency on an unstable supply chain is perceived as an obstacle to effective schistosomiasis control.

In this context, participants stressed the necessity of shifting towards a community-led model, citing precedent in historical public health initiatives.

> “We had other teams in all the facilities for malaria, and we just decided people need to get tested before they get treated. We need to get to that stage with schistosomiasis control, especially when we have limited resources and drugs. So [test and treat] is something there are advantages about.” (KII 4)

Rather than positioning targeted diagnostics as an added intervention, stakeholders frame it as a necessary adaptation to the systemic constraints. Together, these perspectives illustrate a tension for change characterized by erosion of confidence in existing MDA-dependent models and increasing openness to alternative, locally actionable approaches to schistosomiasis control.

Mission alignment between the stakeholders and the toolkit as an intervention in this context is further perceived as a facilitator. CHEWs demonstrated openness to adopting new tools, underscoring that the tension for change is felt at both the systemic and individual levels.

> “If you see any tools at all, we can make use of them…If we are trained on how to use it, we will use it.” (CHEW 1)

Rather than viewing the toolkit as outside of their scope of practice, CHEWs view it as a natural extension of their existing responsibilities. This alignment with their professional identity and commitments reduces anticipated resistance to adoption.

In addition to role congruence, compatibility of the toolkit within stakeholder workflows was largely seen as a facilitator. Again, participants cite precedent in successful shifts towards community-based test and treat models for other public health initiatives, positioning the toolkit as a natural addition rather than fundamentally new or disruptive.

> “It is highly welcomed. We have been doing something similar to that before. It is similar with malaria diagnosis. And we do HIV diagnosis with a toolkit. We do that in a health facility, which are also done by the CHEWs and the nurses. Everyone who is also knowledgeable can do it. We have already seen that workflow in the health facility.” (KII 2)

> “It will [fit into the workflow] because although in the facility we have a lot of work we are doing…if schistosomiasis now comes in, it is part of the examination that we are going to do.” (CHEW 2)

However, compatibility with workflows is potentially impeded by broader constraints related to personnel capacity and workload.

> “The workload on the part of the CHEWs [is a potential challenge] because we already have a shortage of manpower in the healthcare centers… But I also believe that diagnosing with the use of the Foldscope is not something that will be done on a daily basis.” (KII 1)

Finally, both CHEW and KII stakeholders cited a uniquely collaborative relationship between community health providers and the Ministry of Health. Every participant endorsed close collaboration and supervision between CHEWs and the Ministry of Health as a necessary component of the intervention. This distribution of power amongst stakeholders positions discursive power (authority prescribed to the implementer) as a strong facilitator.

> “The supervision should be a supportive supervision, not a type of policing supervision. So you notify [the CHEW] that you are coming for supervision. If they are not doing it fine, you can give them on-the-job training. But I think that’s what you need to do: train the local governments, train the coordinators, and the frontline health facility workers initially.” (KII 1)

Importantly, the emphasis on capacity-building and collaboration frames CHEWs as active participants whose autonomy and competence must be cultivated. Such unanimously endorsed power sharing speaks strongly to a shared sense of trust between stakeholders at individual and systemic levels.

#### INDIVIDUALS: Innovation Recipients

CHEWs cite a high level of trust between themselves and community members, grounded in both their community integration and historical precedent.

> “[The patients] will feel confident…We are the ones that will recommend it…so they will believe us. So far they have confidence in us.” (CHEW 1)

> “They will accept it because it’s like the RDT (rapid diagnostic test) that we normally use to diagnose malaria. It’s also the same thing in this case of schistosomiasis. By the time I bring out my Foldscope and I collect, “Oya go and bring your urine,” they bring it and I am able to do it. Then the community will feel relaxed.” (CHEW 3)

CHEWs appear to benefit heavily from their position as trusted, integrated members of their local communities, leading to broad expectations of acceptability. This trust is particularly crucial for shaping community acceptance and facilitating uptake, reinforcing the central role of community-integrated stakeholders in intervention planning.

##### Panel 1: Implementation determinants identified through Consolidated Framework for Implementation Research (CFIR) qualitative analysis

Consolidated Framework for Implementation Research: Qualitative Assessment of Local Healthcare Stakeholders to assess Feasibility & Acceptability of Toolkit. CFIR constructs presented represent those most salient in participant narratives regarding pre-implementation considerations. CFIR = Consolidated Framework for Implementation Research.

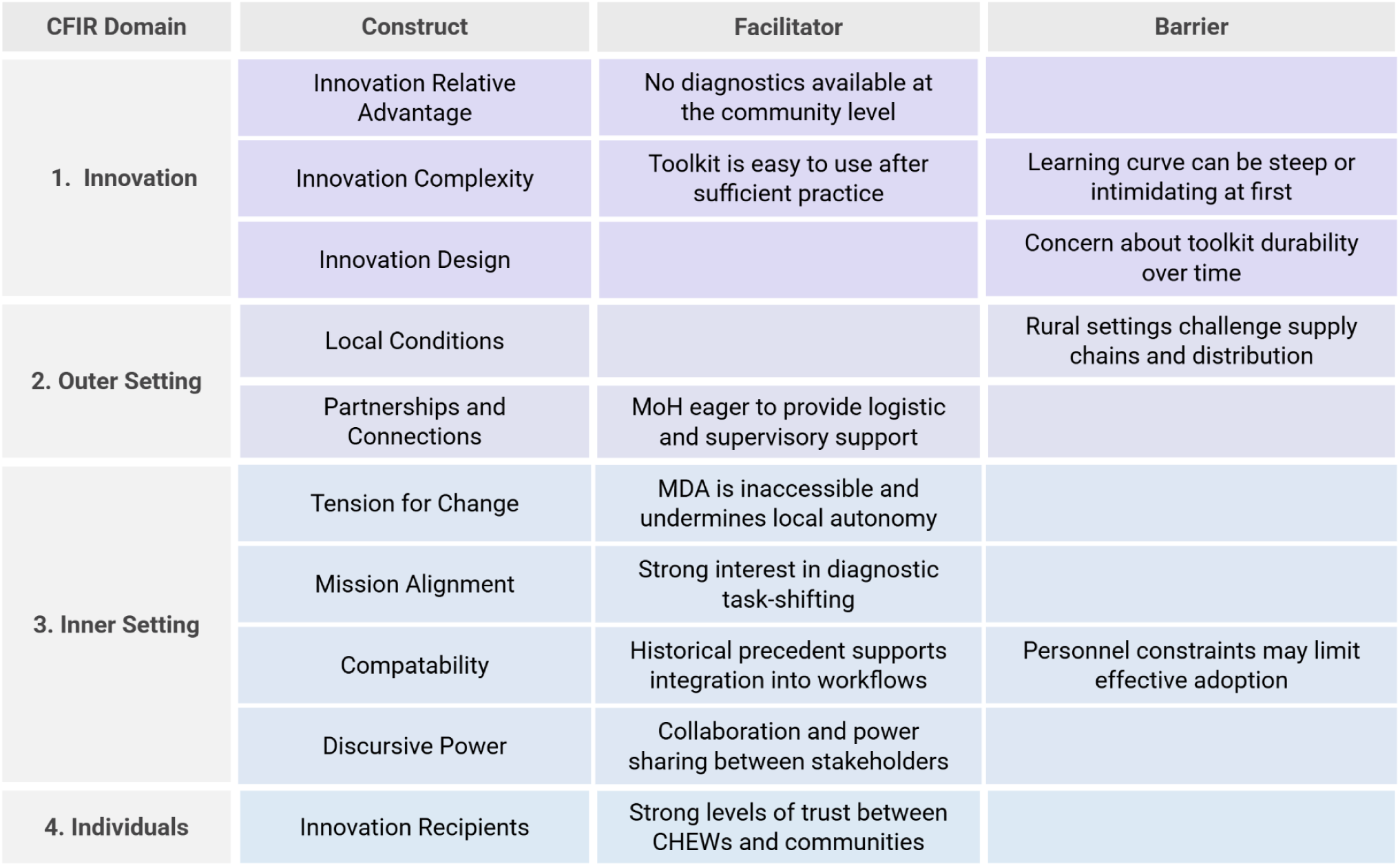

## DISCUSSION

This study provides strong evidence that decentralized, community-led diagnosis of urogenital schistosomiasis is accurate, feasible, and acceptable, supported by quantitative performance metrics and qualitative end-user narratives. First, the Foldscope alone can effectively serve as a substitute for standard microscopy in diagnosing urogenital schistosomiasis, as evidenced by the excellent diagnostic performance (Sensitivity: 90.99%; specificity: 99.34%) of blinded lab scientists using the Foldscopes under laboratory conditions (Supplemental Figure 5). Second, CHEWs, following brief training, progressed from poor initial performance with accuracy of 52.5% (CI: 37.5–67.1) to significantly improved accuracy of 92.1% (79.2–97.3), demonstrating that learning to use the Foldscope-SchistoFilter toolkit (Estimated per-test cost of $0.02-0.04 USD / ₦27-56) is feasible and that diagnostic task-shifting is achievable with appropriate support. This improvement pattern underscores the importance of hands-on training and demonstrates that initial lower performance metrics improve substantially with continued use. The overall pooled diagnostic metrics (Sensitivity: 62.7%; specificity: 74.7%) must be interpreted cautiously, as they mask substantial heterogeneity in performance across the training period and may underestimate post-training operational capability. This learning curve pattern showing significant improvement in accuracy and specificity converges with qualitative narratives from CHEWs who endorsed notable increases in ease-of-use and confidence across subsequent study days.

### POLICY AND PRACTICE IMPLICATIONS

The limitations of MDA in schistosomiasis control warrant immediate attention. Persistent constraints in medication availability and personnel capacity limit the frequency and coverage of effective MDA in many high-risk communities. Furthermore, the limited autonomy of community stakeholders and Ministries of Health in MDA programming, which is reliant on external donors, reiterates the need for community-led approaches. Thus, exploring the role of a test-and-treat model in this context may provide additional options to support schistosomiasis control in communities where effective MDA coverage is difficult to achieve.

The central question is no longer whether a test-and-treat approach is possible but rather how diagnostic capabilities can be integrated into routine community-based practice. By proposing CHEWs as an effective end-user, our findings suggest several complementary implementation strategies through which community-led interventions could be deployed, depending on local epidemiology and health system capacity.

A straightforward entry point for test-and-treat programming is the targeted screening of high-risk populations, including school-aged children and those with consistent exposure to freshwater. Notably, while school-aged children remain a priority population, our study demonstrated a substantial burden among adolescents and young adults, reinforcing the utility of targeted programming outside of traditionally high-risk age groups given the systemic constraints that preclude adult-inclusive MDA.

The second approach involves a test-treat-track strategy that focuses on the targeted screening of all school-aged children and the subsequent tracking and screening of households of all positive cases. Hybrid programs combining MDA and targeted testing may also be effective.

Finally, a symptom-based referral pathway may be most efficacious for communities with very limited personnel capacity. Self-referral for testing requires individuals to recognize the symptoms of schistosomiasis and present themselves to the clinic for testing. However, as demonstrated in Figure 4, a significant gap was identified between those who had heard of schistosomiasis and general symptom awareness beyond hematuria, suggesting that individuals experiencing these symptoms may not associate them with schistosomiasis and therefore may not seek testing. Thus, self-referral without improved symptom awareness is unlikely to result in good diagnostic coverage.

Educational tools targeting symptom awareness have had great success in Oyan River Dam communities in recent years in the form of the “Schisto and Ladders” educational board game (23, 24). Furthermore, the utility of the Foldscope not just as a diagnostic tool but as an educational one (consistent with its original educational design) makes its integration into educational programming a unique benefit. Foldscope-based educational sessions enabling direct parasite visualization could transform abstract concepts of disease into concrete evidence of infection, an approach that has been shown to increase community involvement in UGS control efforts (25).

### FUTURE DIRECTIONS

This study has important implications for diagnostic task-shifting. While CHEWs are important stakeholders in public health interventions, diagnostics have traditionally remained outside of their scope of practice. Our study provides evidence that, with appropriate training and oversight, CHEWs can perform urogenital schistosomiasis diagnosis with high levels of accuracy and confidence, supporting an expanded role in diagnostic endeavors. This work is particularly relevant to areas endemic for schistosomiasis with similar geographic barriers to care, and the authors call for continued study of point-of-care diagnostic strategies, including training of CHEWs in rural areas in Borno, Yobe, Jigawa, Niger, and Lagos states, as well as Kano, Sokoto, Benue states to address the diagnostic gap for these endemic regions (26).

Importantly, possible use-cases of the Foldscope when used by CHEWs extend beyond urogenital schistosomiasis. Previous studies have demonstrated its utility in cervical cancer screening (27) and detection of fungal pathogens in clinical samples (21), and theoretical uses exist for a range of diseases, including *Schistosoma mansonii*, soil-transmitted helminths, leishmaniasis, sickle cell disease, and trichomoniasis (28). Thus, this presents the possibility of cross-training CHEWs, equipping them with a single toolkit capable of screening for multiple diseases that impact their communities. Such approaches are particularly relevant in rural settings where geographic barriers limit access to centralized facilities and multiple neglected tropical diseases co-occur (19,29,30).

The diagnostic toolkit used in the current study represents a “frugal science” approach that does not sacrifice functionality for affordability, creating new opportunities for decentralized health system implementation (31). This frugal science approach may be complemented by machine learning. Paired with the Foldscope’s mobile phone compatibility, one could further enhance diagnostic accuracy to supplement the human eye with automated image detection (32,33). However, such approaches require careful co-development and contextualization to ensure that the intervention is acceptable, useful, and properly situated within social, structural, and pragmatic constraints (34–37).

### PARALLEL COMMUNITY INITIATIVES

This study is one component of a broader, community-centered response to schistosomiasis in Ogun State developed in parallel with the diagnostic work reported here. This work is in alignment with both the Declaration of Helsinki’s 2024 mandate for meaningful community engagement and the WHO NTD 2030 Roadmap’s call for integrated, multi-pronged elimination strategies. Alongside the diagnostic toolkit, the research team has co-developed a microscopy-based educational game and disease ecology-based visualization curriculum for schoolchildren, a population this study identifies as carrying a high burden of infection. Environmental monitoring efforts are also underway, using drone-based remote sensing and community-mediated site selection to map snail habitat along the Oyan River Dam, informing targeted snail control interventions and longitudinal transmission surveillance. Taken together, these initiatives reflect a conviction that durable schistosomiasis elimination requires simultaneous action on diagnosis, education, and the ecological conditions that sustain transmission.

### LIMITATIONS

This study has several limitations. First, the number of samples processed by CHEWs (N=237) did not meet the target sample size (N=365), potentially limiting precision in some performance estimates. Additionally, CHEW performance was evaluated within a supported study environment, and diagnostic accuracy may differ under routine programmatic conditions. Individual, site, and community-level variations may contribute noise to the learning curve data, though schoolhouse clinics were run in similar contexts, conditions were not identical each day, and individuals may learn at different speeds. Learning retention and duration of these skills must also be evaluated. Finally, though this study demonstrates the toolkit’s efficacy, key barriers to implementation, notably perceived toolkit durability and supply chain reliability, must be addressed in any successful implementation effort.

## Data Availability

In accordance with the data sharing provisions of the Ogun State Health Research Ethics Committee (OGHREC/467/2025/618/APP) and UTHealth Houston Institutional Review Board (HSC-MS-25-0502), deidentified datasets will be shared following ethical guidelines and data privacy regulations. The authors intend to deposit deidentified diagnostic data, the study protocol, semi-structured interview guides, and SchistoFilter construction instructions in a publicly accessible repository (Open Science Framework) prior to publication the corresponding DOI will be included in the published article. Raw diagnostic assessment data, template data collection instruments, and a data dictionary will be made available to enable replication of study methods. SchistoFilter device construction instructions and performance data will be made available to support recreation and validation of this open-source sample preparation approach. Individual qualitative interview transcripts will not be shared to protect participant confidentiality aggregated CFIR construct mapping and thematic coding data may be shared upon request, and the semi-structured interview guides will be made available. The study protocol and English and Yoruba versions of informed consent forms, demographic questionnaires, and interview guides will be available six months after publication and upon direct request thereafter. Requests should be directed to.

https://dx.doi.org/10.17504/protocols.io.4r3l2dpkjg1y/v1

https://www.protocols.io/private/1914F3331F4911F19C7D0A58A9FEAC02

## ACKNOWLEDGEMENTS

We thank the community members, leaders, teachers, and students of Imala Odo, Ibaro, Apojola, Abule Titun, and Abule Sikuru for their participation and trust. We acknowledge the staff of the Ogun State Ministry of Health, the Federal Medical Center Abeokuta, and the Ogun State NTD Control Office — particularly the former Director of Public Health, Dr. Festus Soyinka — for institutional support and encouragement. We thank members of the Prakash Lab at Stanford University for contributions to study proceedings. Fixed *S. haematobium* (Egyptian) eggs used for laboratory imaging were provided by the Schistosomiasis Resource Center of the Biomedical Research Institute (Rockville, MD) through NIH-NIAID Contract HHSN272201700014I. Field photography was conducted by Daniel Amao and Thomas Copeland. We thank Dr. Charles Darkoh and Dr. Lu-Yu Huang for their support of the medical students involved, and the volunteers of Health In Your Hands Diagnostics for their contributions.

## CONTRIBUTORS

W.C.M., C.J.C., T.O., and I.A.G. contributed equally as co-first authors and were responsible for conceptualization, study design, methodology, field data collection, statistical analysis, interpretation, and original draft preparation. W.C.M., C.J.C., T.O., H.M., U.F.E., M.A.A., A.O., S.L.B., J.B., R.K., A.K., V.S. and C.S. contributed to protocol design and development. U.F.E. and S.I. led site selection. W.C.M. and N.S. performed formal statistical analyses. S.L.B. and N.S. contributed to quantitative data analysis and data management. C.J.C. led qualitative analyses with contributions from W.C.M., S.L.B., K.N., and H.M. T.O., W.C.M., I.A.G., S.I., and M.K.E. oversaw field study proceedings. H.T.L. led primary data collection and fieldwork, with assistance from V.S. and W.C.M.. W.C.M. and R.K. led participant consenting, with assistance from V.F., M.K.E., and O.B.O. M.H.S. contributed to manuscript writing and geographic mapping. S.B.S., C.S., and J.L. contributed to writing and editing. K.T. and O.A. provided supervisory oversight and contributed to writing, review, and editing. M.P. and J.L. provided senior supervisory oversight, facilitated laboratory resources, and supported all study phases. K.N., O.B.B., P.J.K., M.K.N., O.B.O., I.E., A.E.H., V.F., B.D.O., C.C., F.O., M.R., A.A., A.J.S., A.Ab., K.Na., S.T.A., T.C., and D.A. contributed to data collection. All authors critically reviewed and approved the final manuscript. W.C.M. and M.P. serve as co-corresponding authors.

## DECLARATION OF INTERESTS

Manu Prakash is a co-founder of Foldscope Instruments, Inc., which manufactures the Foldscope microscopes used in this study. All other authors declare no competing interests. Health In Your Hands Diagnostics 501(c)(3) developed the SchistoFilter independently for open-source construction and use by any party. Dr. Prakash’s involvement was driven by scientific and public health objectives. Health In Your Hands Diagnostics 501(c)(3) provided organizational support and facilitated study operations. All other authors declare no competing interests.

## DATA AVAILABILITY STATEMENT

In accordance with the data sharing provisions of the Ogun State Health Research Ethics Committee (OGHREC/467/2025/618/APP) and UTHealth Houston Institutional Review Board (HSC-MS-25-0502), deidentified datasets will be shared following ethical guidelines and data privacy regulations. The authors intend to deposit deidentified diagnostic data, the study protocol, semi-structured interview guides, and SchistoFilter construction instructions in a publicly accessible repository (Open Science Framework) prior to publication; the corresponding DOI will be included in the published article. Raw diagnostic assessment data, template data collection instruments, and a data dictionary will be made available to enable replication of study methods. SchistoFilter device construction instructions and performance data will be made available to support recreation and validation of this open-source sample preparation approach. Individual qualitative interview transcripts will not be shared to protect participant confidentiality; aggregated CFIR construct mapping and thematic coding data may be shared upon request, and the semi-structured interview guides will be made available. The study protocol and English and Yoruba versions of informed consent forms, demographic questionnaires, and interview guides will be available six months after publication and upon direct request thereafter. Requests should be directed to.

## FUNDING

This study was supported by the Gates Foundation (M.P.), Moore Foundation (M.P.), the UTHealth Houston Office of Global Health Initiatives (J.L.), and a crowdfunding campaign hosted on Experiment.com (W.C.M.). Funders had no role in study design, data collection, analysis, interpretation, or the decision to submit for publication.

## AI DISCLOSURE

During manuscript preparation, the authors used Grammarly for grammar checking and Claude AI to review formatting and reference numbering. BioRender’s AI-assisted “Apply BioRender Style” function was used to generate representative cartoons of the toolkit from the study team’s photography. All AI-assisted outputs were reviewed and edited by the authors, who take full responsibility for the published content. No AI language models were used to generate data, conduct analyses, or interpret results. All authors are responsible for the accuracy and integrity of the reported findings.

## SUPPLEMENTARY APPENDIX

SUPPLEMENTARY NOTE 1 | Ethical considerations

Local, Ogun State, and U.S.-based ethical approval was sought and obtained prior to study initiation. Verbal approval from village leaders was enabled by community sensitization visits conducted with oversight from the Ogun State Ministry of Health NTD Office. The Ogun State Health Research Ethics Committee approved the study on June 25, 2025 (OGHREC/467/2025/618/APP). UTHealth Houston’s Institutional Review Board and Committee for Protection of Human Subjects approved all study proceedings, consent forms, and personnel roles (IRB # HSC-MS-25-0502).

SUPPLEMENTARY NOTE 2 | The Foldscope-SchistoFilter toolkit: an alternative to the gold standard

Critical to the practical implementation of any community-based diagnostic approach to schistosomiasis is the development of sample preparation methods that do not require expensive consumables, laboratory infrastructure, or specialized personnel [1,2]. Traditional urine concentration techniques rely on polycarbonate filters or centrifugation, requiring electricity or single-use laboratory consumables. The Sterlitech polycarbonate filter is currently favored for field diagnostic efforts due to its reliable ability to concentrate eggs from urine, but its cost and lack of reusability make it impractical in rural settings.

The Foldscope is a commercially available, electricity-free, paper-based microscope with a unit cost under $2.50 USD. An initial proof-of-concept study using the Foldscope for UGS diagnosis in Ghana identified the need for simplified filtration methods and validation by non-expert users [3]. Since that study, substantial improvements in both Foldscope design and smartphone capabilities have enhanced the feasibility of field-based microscopy. The Foldscope v2.0 now offers 340× magnification (up from 140×), and advances in smartphone camera quality enable higher-resolution imaging through the optional smartphone attachment. Increasing availability of high-resolution smartphones in low- and middle-income countries makes reliable image capture by community health workers like CHEWs increasingly feasible.

Diagnostic testing — particularly tasks requiring microscopic evaluation — falls outside the current scope of CHEW daily practice. This study therefore employed a mixed-methods design to evaluate not only diagnostic performance, but also the perspectives and lived experiences of CHEWs regarding the intervention, and the broader feasibility of task-shifting urine sample evaluation to community health workers. Successful implementation of any community-based diagnostic program requires understanding and addressing the concerns of the communities and end-users it is intended to serve.

## SUPPLEMENTARY METHODS

### SCHISTOFILTER CONSTRUCTION AND USE

The SchistoFilter consists of four primary components in a layered configuration. The coverslip layer (60 mm × 50 mm) was fabricated from 0.25 mm thick clear PETG sheet (McMaster-Carr, part #4076N11). The base layer (65 mm × 55 mm) was cut from 0.5 mm thick clear, scratch- and UV-resistant acrylic film (McMaster-Carr, part #85815K211) with a 5 mm diameter aperture positioned centrally.

The filtration membrane was constructed by excising an 8 mm diameter circle from a 38 mm circular stainless steel filter gasket containing 550-mesh (25-micron pore size) 304 stainless steel screen (USA Lab). The circular mesh segment was affixed to the base layer using all-purpose contact adhesive (Seal-All), positioned to completely occlude the 5 mm aperture, and cured for 8–12 hours at room temperature.

To complete assembly, a 65 mm × 55 mm vinyl adhesive backing layer (Cricut Smart Vinyl, permanent) with a central 5 mm aperture was applied to the reverse side of the base, creating a sandwich configuration with the metal mesh. This produced a 3 mm overlap between the 8 mm mesh perimeter and the 5 mm vinyl aperture on all edges, securing the filtration membrane and creating a sealed sample chamber. The device can also be constructed from basic supplies with scissors or a razor blade, laminate sheets, and a hole punch, using any stiff backing card, so long as the final thickness is between 0.7 mm and 1.7 mm for optimal focus.

To use: draw urine into a syringe and expel slowly through the filtration membrane, trapping Schistosoma eggs. Secure the coverslip with paperclips and assemble into the Foldscope by pulling down the backflap and inserting the sample into the stage. Position a phone (via Foldscope magnetic pairing ring) or eye over the lens to view. To reuse, remove the SchistoFilter from the Foldscope, wash the filtration membrane with three syringe flushes of water from the reverse side of the central aperture, and dry with a paper towel or alcohol wipe.

### DEVELOPMENT AND BENCH VALIDATION OF THE SCHISTOFILTER

We developed the SchistoFilter as a point-of-care filtration device to concentrate Schistosoma eggs from urine without requiring electricity or laboratory consumables. Bench validation used schistosome eggs previously isolated from rodent models of schistosomiasis. By suspending 50–200 eggs in 10 mL of water, drawing the liquid into a syringe, and pushing the fluid through the membrane, the team was consistently able to image trapped eggs on the SchistoFilter’s stainless steel mesh using both a confocal microscope (Revolve ECHO) and a Foldscope, confirming the device’s efficacy across a range of egg concentrations.

### SAMPLE SIZE CALCULATION

Sample size was calculated to estimate sensitivity and specificity within ±5 percentage points at 95% confidence. Based on a 52% prevalence estimate from prior epidemiological surveys in Oyan River communities [2,5,19] and an assumed sensitivity of 80%, the standard diagnostic accuracy formula indicated 473 participants were needed. Applying a finite population correction for the study communities (population ≈1,600), the adjusted target was 365 participants across five communities. We recruited 418 participants to account for potential dropout; 237 completed the full diagnostic protocol including CHEW point-of-care assessment.

The standard formula applied was: N = [Z² × Se × (1 − Se)] / [d² × P], where N is the minimum required number of infected individuals, Z is the standard normal deviate at 95% confidence (1.96), Se is the expected sensitivity (0.80), d is the desired precision (0.05), and P is the estimated prevalence (0.52). This yielded a target of 365 participants.

### ADDITIONAL PARTICIPANTS AND CHEW RECRUITMENT

Following documented verbal consent, CHEWs participated in eight days of combined training and data collection. Six CHEWs participated for all eight days; one began on day four and another withdrew after day four due to external commitments.

Laboratory scientists recruited from the Federal University of Agriculture, Abeokuta, and the Ministry of Health conducted reference standard microscopy at field sites. Following field data collection, one participating laboratory scientist trained by the research team subsequently cascade-trained a team of students to carry out blinded secondary analysis of stored field samples using the Foldscope with gold standard (SterliTech) filtration.

The research team collaborated closely with officials from the local and state Ministries of Health throughout. Recruited members from the Department of Neglected Tropical Disease provided clinical oversight and assisted with community sensitization but were not directly involved in data collection or analysis. At the conclusion of the study, these individuals were recruited and consented as key informant interview participants.

### PARTICIPANT COMPENSATION

Community member participants did not receive the results of their reference standard screening; however, 404/418 participants were administered praziquantel in an age- and height-appropriate dose by the resident Ministry of Health doctor as preventive chemotherapy, in line with MDA protocol. Community members also received approximately $1.50 USD (₦2,000) and a pack of snacks and beverages for their participation. Eight CHEWs received a total of $140 USD (₦200,000). Laboratory scientists were compensated $229 USD (₦320,000). Ministry of Health counterparts received between $140–229 USD (₦200,000–320,000) for their clinical oversight, assistance, and interview participation.

### TRAINING OF THE CHEWS

Training sessions emphasized technical use of the tools and morphological identification of parasite eggs by size, color, and shape. CHEWs were considered ready to assess clinical samples once they could successfully identify eggs on glass slides and correctly operate each Foldscope component (focus ramp, light module orientation, and slide loading). Urine filtering and SchistoFilter assembly were demonstrated with a clinical sample aliquot before the study period began. Sample size for the CHEW validation subset is limited to n=237 due to the timing and practical constraints of operating within a small schoolhouse transient clinic.

### DETAILED SAMPLE COLLECTION AND PREPARATION

Urine samples were collected at midday (10:00 –14:00). Each participant provided a minimum of 30 mL of urine into a sterile container (CLINSAM 90 mL sterile urine collection cups). Each sample then underwent two parallel filtration processes: (1) conventional gold standard urine filtration and (2) SchistoFilter filtration.

For the gold standard technique, a 10 mL aliquot was drawn into a Luer-lock syringe (BH Supplies BH110L) and expelled through a 20 µm polycarbonate membrane filter (Sterlitech) in a filter holder (Sterlitech). The membrane was placed on a glass slide and examined under a compound microscope (AmScope B120, 10–40× objectives) by an Ogun State Ministry of Health NTD laboratory scientist. The whole slide was scanned and total egg counts recorded as eggs per 10 mL urine on a de-identified paper chart. Any sample with at least one identified S. haematobium egg was classified positive; samples with no identified eggs were classified negative. After counting, a drop of 10% formalin was added to each filter, covered with a glass coverslip, and sealed with clear nail polish for long-term ambient storage.

For the SchistoFilter condition, a separate 10 mL aliquot from the same collection cup was expelled through the SchistoFilter membrane by a trained CHEW. The filter was inserted into a Foldscope and examined via a Tecno Pop 10 smartphone attachment. Egg counts were recorded using the same classification criteria. After visualization, the SchistoFilter was washed in two sequential bowls of sterile sachet water, dried on clean paper towels, and prepared for reuse.

### FOLDSCOPE-BASED SMARTPHONE IMAGING

During a preliminary meeting in Abeokuta with Ogun State clinicians, NTD coordinators, and research collaborators, the team evaluated several locally available smartphones to identify a cost-effective option for image capture. The Tecno Pop 10, widely owned by collaborators, provided the best balance of camera quality and affordability. Two used Tecno Pop 10 phones were purchased from a local marketplace for field use. Tape was applied to each phone to secure the Foldscope magnetic pairing ring and ensure stable alignment.

### STANDARD MICROSCOPY IMAGES

Lab-based images of *S. haematobium* eggs were acquired using a Revolve ECHO fluorescence microscope with dual 12MP CMOS color and 5MP CMOS mono cameras (20× and 40× objectives). Field photographs of gold standard microscopy were captured with an AmScope B120 LED Binocular Compound Microscope (10× objective). Scale bars were formatted using Apple Numbers.

### QUALITATIVE CFIR ANALYSIS

Interviews were audio recorded via SurveyCTO and manually transcribed verbatim by research team members. Unclear audio segments were reviewed by a second team member. Transcripts were uploaded into Dedoose for coding and thematic analysis. A primary coder coded all transcripts; a 25% subset was independently coded by two additional coders. Subsequent analysis of barriers and facilitators used the Consolidated Framework for Implementation Research (CFIR) collaboratively across coders.

Coding applied a combined deductive and inductive framework. Deductive codes were derived from the 48 CFIR constructs spanning five domains: innovation, outer setting, inner setting, individuals (roles and characteristics), and implementation process. Inductive codes captured themes emerging from the data not covered by existing CFIR constructs. See Supplementary Table 7 for the COREQ 32-item checklist.

### DATA MANAGEMENT

A total of 418 participants were recruited across five communities. SurveyCTO digital forms were used for demographic data collection and consenting; electronically signed English and Yoruba consent forms were stored on a password-protected digital drive. Complete demographic data were available for 366/418 (87.6%) participants. Missing data arose from software synchronization difficulties (n=52) during days one and two, and were unrelated to participant characteristics or diagnostic outcomes. All demographic analyses use the 366 participants with complete data; diagnostic accuracy analyses include all participants with testable samples. See Supplementary STROBE Diagram.

Field data collection sheets were scanned, transcribed, and uploaded to a spreadsheet aligned with SurveyCTO demographic exports. Each response was verified by participant ID in a linking log. For diagnostic accuracy results, all collected data (n=418) were used for the relevant variables (positive/negative classification, time, egg count).

### STATISTICAL ANALYSIS

Quantitative data were analyzed using GraphPad Prism version 10.5 (GraphPad Software, San Diego, CA) and R version 4.5.2 (R Foundation for Statistical Computing) via Posit Cloud. Statistical significance was set at p<0.05. See end-of-appendix tables for full statistical test details.

Prevalence and demographic analyses: Prevalence and proportions were calculated with 95% Wilson score confidence intervals. Chi-square tests evaluated associations between categorical variables and infection status. One-way ANOVA compared continuous variables across groups, with Tukey’s HSD post-hoc test for multiple comparisons. The correlation between age and infection intensity was assessed using Pearson’s r.

Diagnostic accuracy metrics: Sensitivity, specificity, PPV, NPV, and overall accuracy were calculated from 2×2 contingency tables against the WHO-approved Sterlitech reference standard, with 95% Wilson score confidence intervals. Cohen’s kappa (κ) was calculated with the following interpretation: κ<0.20 (poor), 0.21–0.40 (fair), 0.41–0.60 (moderate), 0.61–0.80 (substantial), 0.81–1.00 (almost perfect). Fisher’s exact test (two-tailed) assessed significance of association between index test and reference standard. Positive and negative likelihood ratios were calculated to assess clinical utility (LR+ >10 and LR− <0.1 indicate strong diagnostic value).

CHEW learning curve: Changes in CHEW diagnostic performance across five sequential communities were assessed using the Cochran-Armitage trend test, a non-parametric test for monotonic trends in binary outcomes across ordered groups. For each metric (accuracy, sensitivity, specificity), data were organized into 5×2 contingency tables. Linear regression was additionally performed for each metric to characterize improvement trajectories. Cumulative error rates were calculated as the proportion of incorrect diagnoses among all samples examined to that point. The relationship between sample number and error rate was assessed using Spearman’s ρ.

Laboratory technician Foldscope validation: Diagnostic accuracy metrics were calculated as above (n=412). Time-to-assessment data were non-normally distributed (p<0.001 by normality testing); differences between positive and negative samples were assessed using the Mann-Whitney U test (two-tailed). The relationship between sequential sample number and time-to-assessment was examined by linear regression.

### CHEW DIAGNOSTIC ACCURACY SUMMARY

|  | Value | 95% CI |
| --- | --- | --- |
| Test | Fisher's exact test |  |
| P value | <0.0001 |  |
| P value summary | **** |  |
| One- or two-sided | Two-sided |  |
| Statistically significant (P<0.05)? | Yes |  |
| Sensitivity | 0.6267 | 0.5135 to 0.7274 |
| Specificity | 0.7469 | 0.6748 to 0.8076 |
| Positive Predictive Value | 0.5341 | 0.4306 to 0.6347 |
| Negative Predictive Value | 0.8121 | 0.7418 to 0.8667 |
| Likelihood Ratio | 2.476 |  |
| CI method | Wilson-Brown |  |

LABORATORY SCIENTIST DIAGNOSTIC ACCURACY SUMMARY
|  | **Value** | **95% CI** |
| --- | --- | --- |
| Test | Fisher's exact test |  |
| P value | <0.0001 |  |
| P value summary | **** |  |
| One- or two-sided | Two-sided |  |
| Statistically significant (P<0.05)? | Yes |  |
| Sensitivity | 0.9099 | 0.8421 to 0.9503 |
| Specificity | 0.9934 | 0.9761 to 0.9988 |
| Positive Predictive Value | 0.9806 | 0.9319 to 0.9965 |
| Negative Predictive Value | 0.9676 | 0.9415 to 0.9823 |
| Likelihood Ratio | 136.9 |  |
| CI method | Wilson-Brown |  |

Following field data collection, 412 archived urine filtration slides that remained undamaged during transport underwent blinded re-examination by a team of six laboratory technicians at the Ogun State Ministry of Health and Federal University of Agriculture, Abeokuta. Examiners were biology, biochemistry, botany, public health, and nursing students with no prior Foldscope training; each received a single training session from one member of the research team. Slides were labeled only by sample identification number; gold standard results were stored digitally by the UTHealth Houston team. Each slide was examined independently using a Foldscope. Time-to-diagnosis was recorded for each sample.

Fisher’s exact test demonstrated significant agreement between methods (p<0.0001, two-sided). Cohen’s kappa was 0.918, indicating excellent agreement with conventional microscopy. Time-to-assessment showed high initial variability (range 5–20 minutes) that progressively decreased and stabilized to 3–8 minutes after 100 samples, with median time declining from approximately 10 to 5 minutes as examiners gained proficiency (p<0.0001, linear regression).

## SUPPLEMENTARY RESULTS

### POOLED DIAGNOSTIC ACCURACY OF CHEWS

Across all 237 assessments, CHEWs demonstrated pooled sensitivity of 62.7% (95% CI 51.4–72.7%), specificity of 74.7% (67.5–80.8%), and overall accuracy of 70.9% (64.8–76.3%). Fisher’s exact test confirmed a significant association between CHEW assessments and reference standard microscopy (p<0.001), with fair overall agreement (Cohen’s κ=0.357). These pooled metrics reflect performance across the full training arc and should be interpreted alongside the sequential community data presented in the main text, which demonstrate marked improvement over time.

## SUPPLEMENTARY DISCUSSION

### LABORATORY VALIDATION AND POTENTIAL TO TRANSFORM DIAGNOSTICS

Our laboratory-based performance builds upon preliminary studies demonstrating Foldscope’s capacity for UGS diagnosis [3]. Laboratory scientists achieved sensitivity of 90.99% and specificity of 99.34%, with accuracy plateauing at 95% after approximately 100 samples — establishing Foldscope microscopy as technically capable of replacing conventional compound microscopy for *S. haematobium* detection. For the purposes of sustainable schistosomiasis control, however, these authors believe the more consequential finding is the feasibility of CHEW-operated diagnosis, which addresses the diagnostic gap at the community level where it matters most.

### DEMOCRATIZING DIAGNOSTICS: FEASIBILITY BY NON-SPECIALISTS

Our study provides strong evidence that microscopy — traditionally considered a specialized laboratory skill — can be successfully performed by CHEWs with appropriate tools and brief structured training. The significant learning curve documented by Cochran-Armitage trend analysis (accuracy: Z=3.08, p=0.002; specificity: Z=3.00, p=0.003) demonstrates that community health workers can achieve high diagnostic accuracy across sequential deployments. The Foldscope’s simple optics, robust construction, and integrated smartphone imaging democratize a technique previously restricted to laboratory settings, enabling rapid capacity building without dependence on expert trainers. Qualitative data emphasizing ongoing supervision needs underscore that initial training must be coupled with quality assurance mechanisms to sustain performance.

### CLINICAL UTILITY ACROSS EPIDEMIOLOGICAL CONTEXTS

In high-prevalence settings (>25%, as in Ibaro and Apojola), even initial moderate sensitivity of 60% provides acceptable positive predictive value (55% in our study), supporting Foldscope use for mass screening campaigns. In low-prevalence settings approaching elimination (<10%), the post-learning-curve performance (82% sensitivity, 93% specificity) and high negative predictive value (97%) validate Foldscope’s role in confirmation-of-elimination surveys. The 11-minute turnaround time enables same-day treatment decisions — addressing the major limitation of conventional approaches that require sample transport with 24–48 hour result delays.

### DEMOGRAPHIC DATA AND IMPLEMENTATION STRATEGY

Students comprised the largest absolute infected population (61 cases), underscoring the importance of considering both relative risk and population size in program planning. The high burden among adolescents and young adults — groups typically excluded from school-based MDA — reinforces the utility of targeted community-wide programming. Complementary youth-centered education approaches are needed to improve symptom awareness and treatment uptake, particularly given the low complication knowledge (39.9%) identified in this study.

### COMPARISON TO OTHER POINT-OF-CARE APPROACHES

The Foldscope system compares favorably to alternative POC approaches on portability, result speed, and cost. The fully reusable design — the SchistoFilter is washed between patients, the Foldscope requires no consumables, and one SchistoFilter was used at least 100 times in our field study — reduces the per-test cost to an estimated $0.02–0.04 USD, compared to approximately $1.50 for Sterlitech filtration alone. This reusability dramatically lowers the cost floor compared to any existing microscopy solution and removes the consumable supply chain dependence that threatens sustainability in the outer setting.

Emerging molecular diagnostics offer superior sensitivity for low-intensity infections but currently require cold chain, reagent supply chains, and higher per-test costs that challenge field deployment [4]. The Foldscope’s microscopy-based approach additionally enables egg quantification for infection intensity assessment, critical for morbidity monitoring and treatment response evaluation [5]. In lower-intensity infections — the majority of cases in this study — molecular diagnostics could provide complementary value where egg detection is unreliable.

### ECONOMIC AND ECOLOGICAL INTEGRATION

While the Foldscope system reduces per-test costs to $0.02–0.04 USD (₦27–56), persistent transmission requires repeated screening indefinitely. Environmental interventions require higher upfront investment but provide sustained impact without ongoing operational cost. Snail habitat removal trials in Senegal have demonstrated reductions in illness burden alongside agricultural and economic benefits [6]. Native freshwater prawn aquaculture simultaneously reduces snail populations and provides protein and income without disrupting local ecology [7]. Solutions to schistosomiasis must offer multi-factorial, synergistic benefits — the diagnostic kit presented here combines portability, low per-test cost, and community agency through CHEW-led diagnosis, offering one component of an integrated response.

Sustainable schistosomiasis elimination requires addressing both downstream effects through improved diagnostics and upstream causes through environmental management [8,9]. As Nigeria pursues WHO 2030 elimination targets, recognizing schistosomiasis as a social-ecological problem — requiring social-ecological solutions alongside MDA and test-and-treat approaches — will be essential [10].

## SUPPLEMENTARY FIGURES

**Supplementary Fig. S0.**
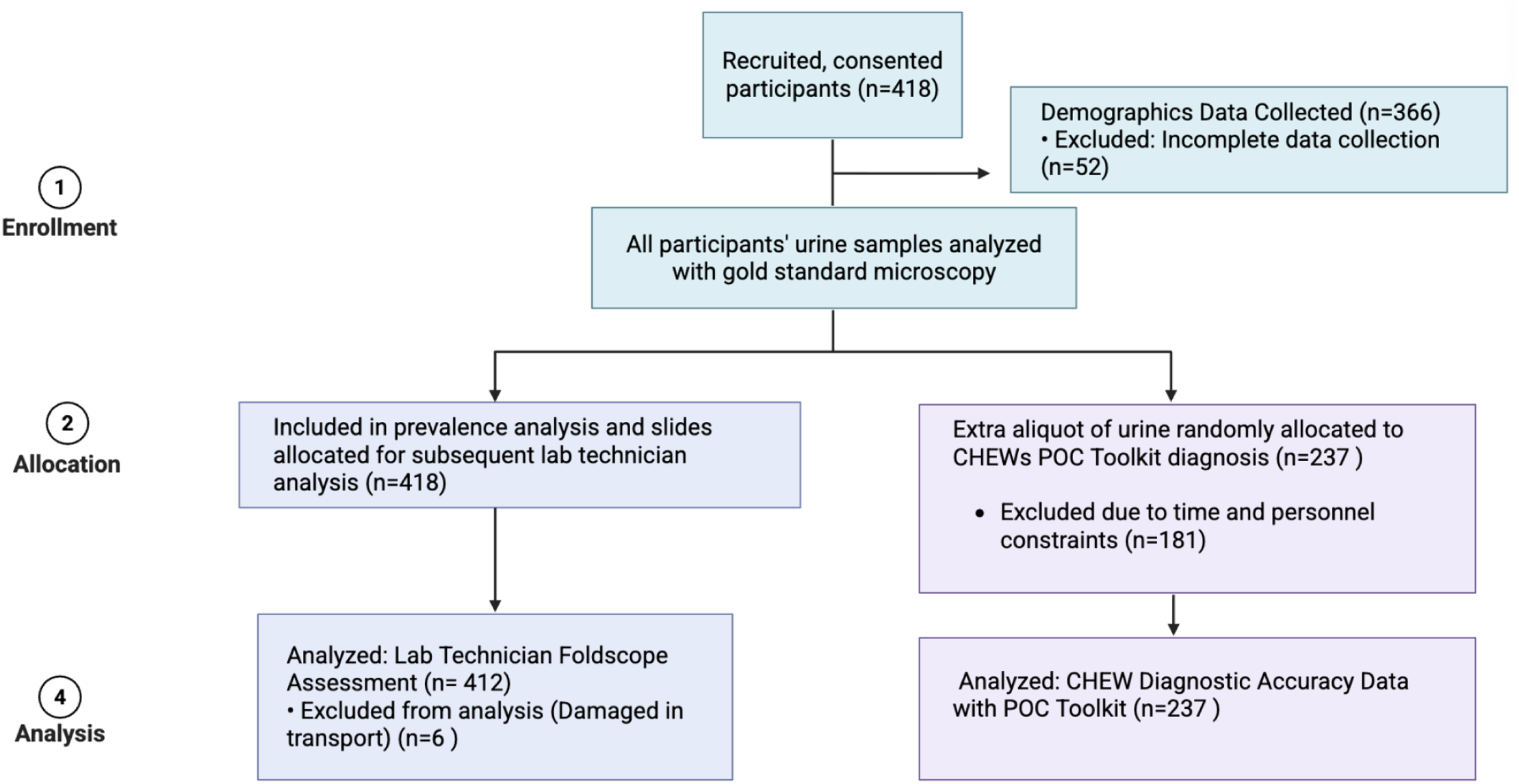
STROBE diagram.

**Supplementary Fig. S1.**
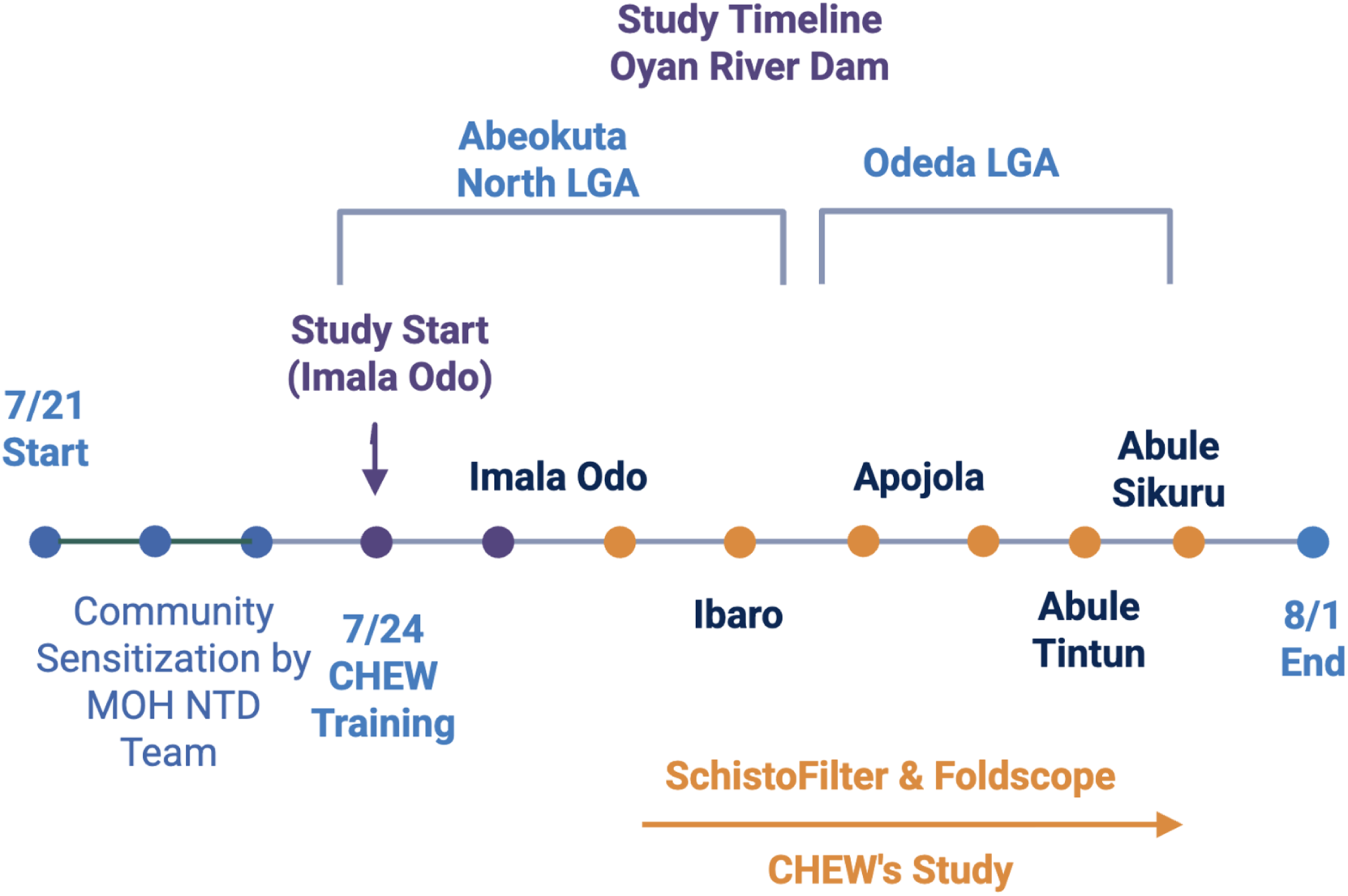
Study timeline. Illustration of the dates and daily activities of the study across eight field days, including training days, sequential community deployments, and key informant interviews.

**Supplementary Fig. S2.**
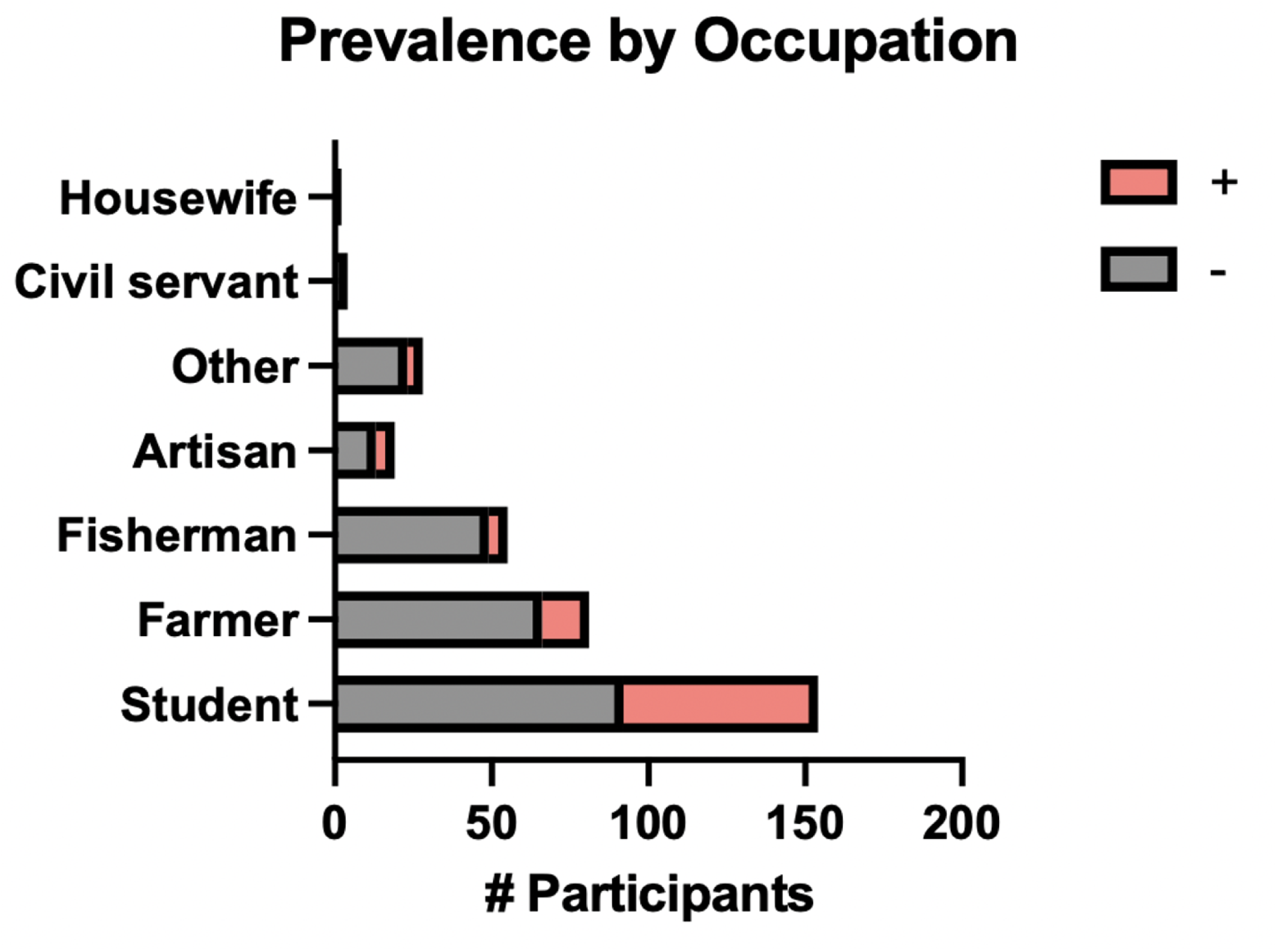
Relative prevalence of urogenital schistosomiasis by reported occupation. Prevalence of S. haematobium infection among community members distributed by self-reported occupation across all five study communities.

**Supplementary Fig. S3.**
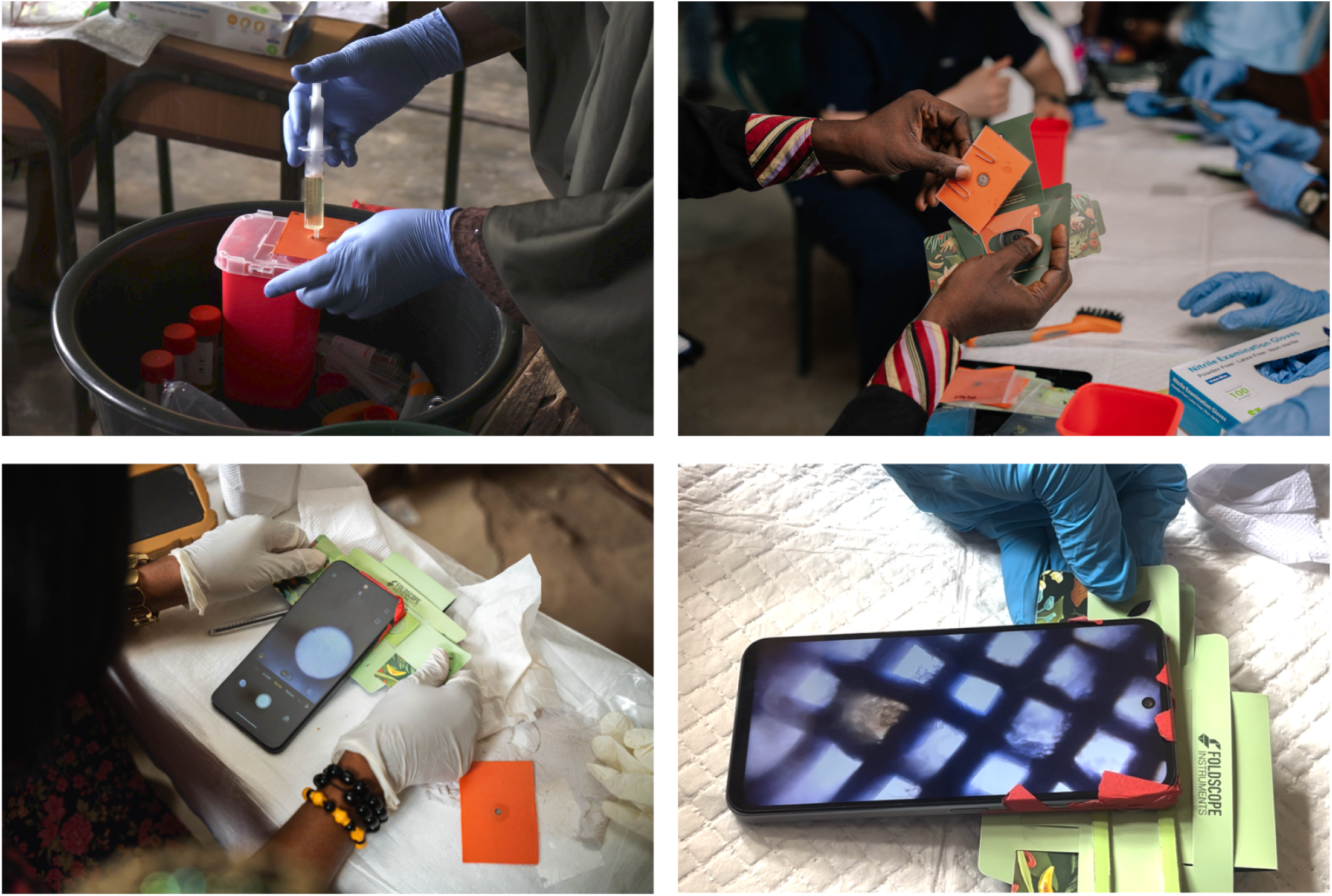
Field photographs of CHEW point-of-care assessment. Community health extension workers (CHEWs) conducting point-of-care sample processing and microscopic examination using the Foldscope-SchistoFilter toolkit in transient schoolhouse clinic settings across study communities.

**Supplementary Fig. S4.**
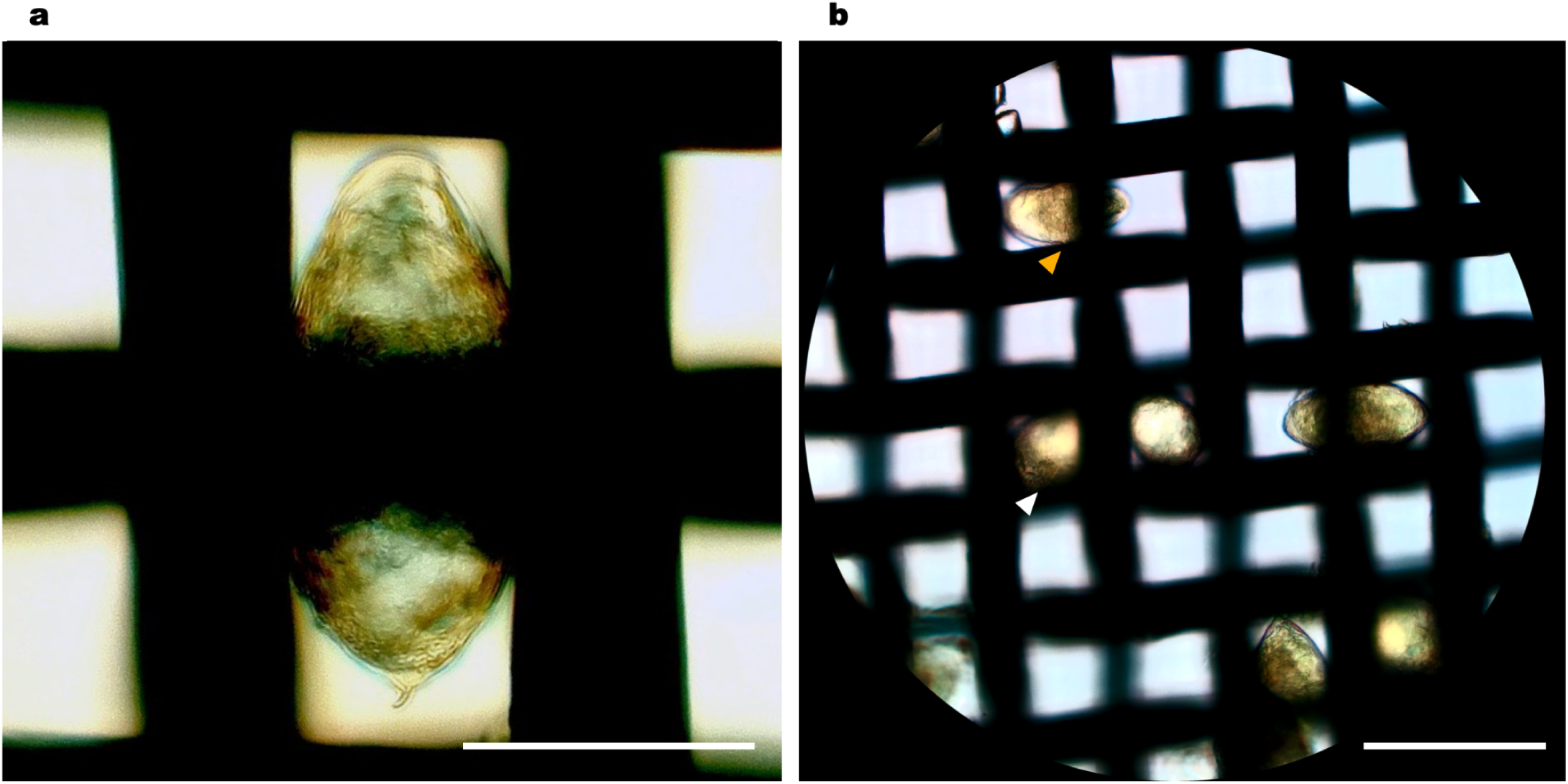
Bench validation of the SchistoFilter. **(a)** *S. haematobium* egg with terminal spine; cropped image at 20× magnification. Scale bar, 65 µm. **(b)** *S. haematobium* ova trapped horizontally (orange arrow) and vertically (white arrow) in the SchistoFilter’s 550-mesh stainless steel membrane (25-micron pore diameter). 20× magnification. Scale bar, 130 µm.

**Supplementary Fig. S5.**
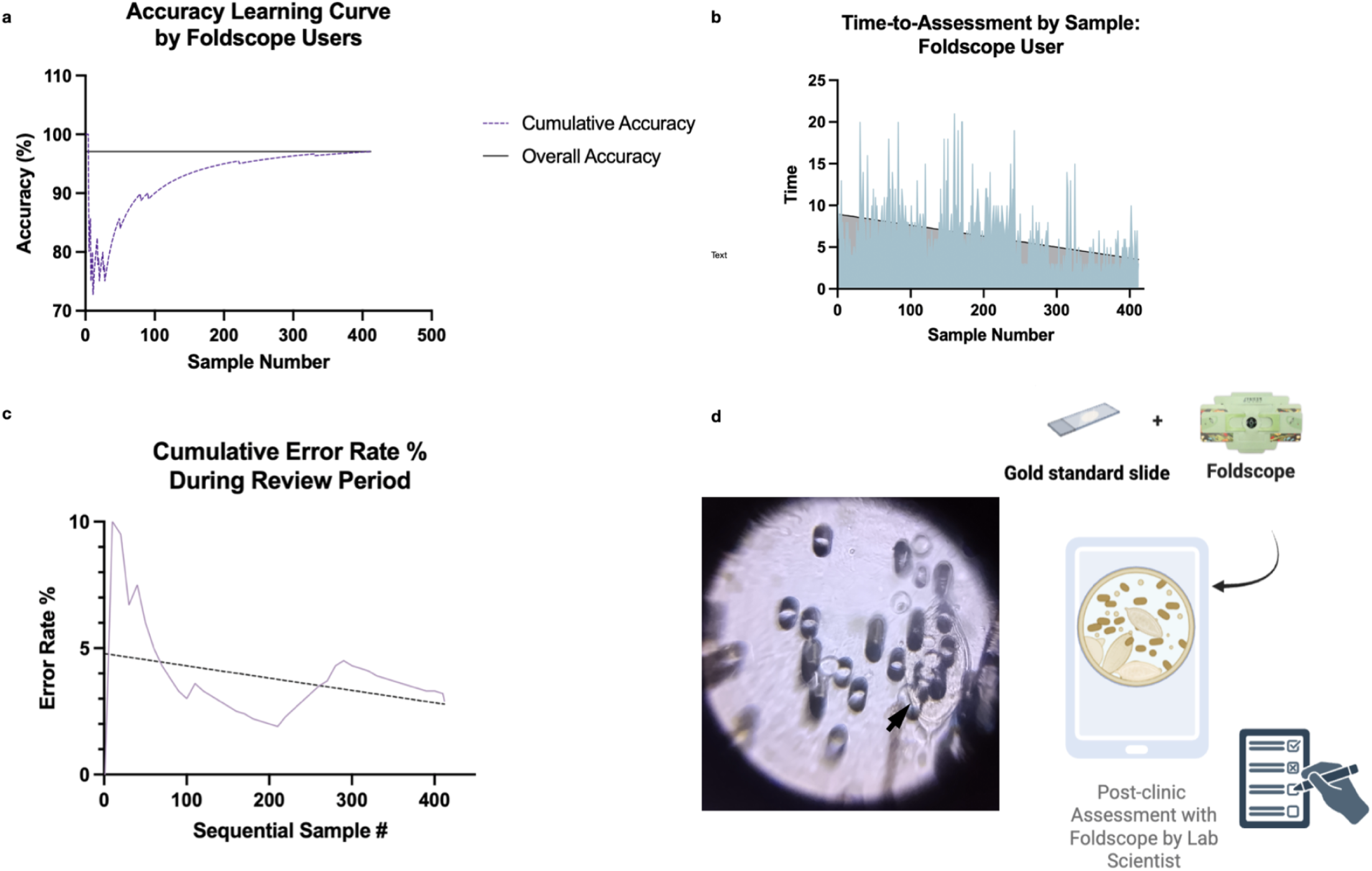
Laboratory validation of Foldscope performance under optimal conditions. Post-study validation demonstrating Foldscope diagnostic accuracy when operated by trained laboratory scientists under controlled conditions. Following field study completion, 412 formalin-preserved reference standard slides (Sterlitech membrane filters) were examined by blinded laboratory scientists using Foldscope microscopy. **(a)** Cumulative (solid line) and rolling 30-sample window (dashed line) accuracy by sequential sample number. Cumulative accuracy stabilized above 95% after ∼100 samples and plateaued at 97–98% by sample 200. Final performance: 97.1% accuracy (95% CI 95.0–98.4%). **(b)** Time-to-assessment per sample showing rapid skill acquisition. Initial high variability (range 5–20 minutes) progressively decreased and stabilized to 3–8 minutes after 100 samples. Median time declined from ∼10 to 5 minutes with experience (p<0.0001, linear regression). **(c)** Cumulative and rolling error rates by sequential sample number, demonstrating progressive improvement in diagnostic precision. Final cumulative error rate: 2.9%. **(d)** Schematic of laboratory technician protocol: formalin-preserved Sterlitech membrane filters on glass slides are inserted into the Foldscope (340×) for examination; the viewer classifies each sample (positive ≥1 egg vs. negative) and documents findings. Representative image taken by newly trained lab scientist, with black arrow pointing to *S. haematobium* ova resting on Sterlitech membrane filter. Overall performance: sensitivity 91.0% (95% CI 84.2–95.0%), specificity 99.3% (95% CI 97.6–99.9%), Cohen’s κ=0.918.

## SUPPLEMENTARY TABLES

**Supplementary Table 1.** Prevalence by community.

| Village | N tested | Positive | Negative | Prevalence (%) |
| --- | --- | --- | --- | --- |
| Imala Odo | 145 | 31 | 114 | 21.4 |
| Ibaro | 65 | 20 | 45 | 30.8 |
| Apojola | 106 | 38 | 68 | 35.8 |
| Abule Tuntun | 52 | 8 | 44 | 15.4 |
| Abule Sikuru | 50 | 18 | 32 | 36.0 |
| Overall | 418 | 115 | 303 | 27.5 |

**Supplementary Table 2.** Prevalence by age group and occupation.

| Characteristic | N (%) | Positive n (%) | Prevalence (%) |
| --- | --- | --- | --- |
| Gender |  |  |  |
| Male | 161 (46.9%) | 49 (30.4%) | 30.4% |
| Female | 182 (53.1%) | 45 (24.7%) | 24.7% |
| Age group (years) |  |  |  |
| 5–14 | 131 (38.2%) | 50 (38.2%) | 38.2% |
| 15–24 | 41 (12.0%) | 22 (53.7%) | 53.7% |
| 25–34 | 46 (13.4%) | 10 (21.7%) | 21.7% |
| 35–44 | 51 (14.9%) | 4 (7.8%) | 7.8% |
| 45–54 | 37 (10.8%) | 5 (13.5%) | 13.5% |
| ≥55 | 37 (10.8%) | 3 (8.1%) | 8.1% |
| Occupation |  |  |  |
| Student | 154 (44.9%) | 62 (40.3%) | 40.3% |
| Farmer | 81 (23.6%) | 15 (18.5%) | 18.5% |
| Fisherman | 55 (16.0%) | 6 (10.9%) | 10.9% |
| Artisan | 19 (5.5%) | 6 (31.6%) | 31.6% |
| Other | 28 (8.2%) | 5 (17.9%) | 17.9% |
| Civil servant | 4 (1.2%) | 0 (0.0%) | 0.0% |
| Housewife | 2 (0.6%) | 0 (0.0%) | 0.0% |

**Supplementary Table 3A.** CHEW toolkit performance relative to reference standard (n=237). Complete confusion matrix for CHEWs’ performance with the point-of-care diagnostic toolkit.

|  | Reference standard<br>(+) | Reference standard<br>(-) | Total |
| --- | --- | --- | --- |
| SchistoFilter + Foldscope<br>(+) | 47 | 41 | 88 |
| SchistoFilter + Foldscope<br>(-) | 28 | 121 | 149 |
| Total | 75 | 162 | 237 |

**Supplementary Table 3B.** Complete CHEW diagnostic performance metrics by community (n=237).

| Sequential<br>Community (L<br>to R) | Imala Odo<br>(n=40) | Ibaro<br>(n=42) | Apojola<br>(n=87) | Abule<br>Titun<br>(n=30) | Abule<br>Sikuru<br>(n=38) | Overall<br>(n=237) |
| --- | --- | --- | --- | --- | --- | --- |
| Prevalence, % | 30.0 | 33.3 | 36.8 | 20.0 | 28.9 | 31.6 |
| Sensitivity, %<br>(95% CI) | 50.0<br>(25.4–74.6) | 78.6<br>(52.4–92.4) | 53.1<br>(36.4–69.1) | 66.7<br>(30.0–90.3) | 81.8<br>(52.3–94.9) | 62.7<br>(51.4–72.7) |
| Specificity, %<br>(95% CI) | 53.6<br>(35.8–70.5) | 75.0<br>(56.6–87.3) | 78.2<br>(65.6–87.1) | 66.7<br>(46.7–82.0) | 96.3<br>(81.7–99.3) | 74.7<br>(67.5–80.8) |
| PPV, % (95% CI) | 31.6<br>(15.4–54.0) | 61.1<br>(38.6–79.7) | 58.6<br>(40.7–74.5) | 33.3<br>(13.8–60.9) | 90.0<br>(59.6–98.2) | 53.4<br>(43.1–63.5) |
| NPV, % (95% CI) | 71.4<br>(50.0–86.2) | 87.5<br>(69.0–95.7) | 74.1<br>(61.6–83.7) | 88.9<br>(67.2–96.9) | 92.9<br>(77.4–98.0) | 81.2<br>(74.2–86.7) |
| Accuracy, %<br>(95% CI) | 52.5<br>(37.5–67.1) | 76.2<br>(61.5–86.5) | 69.0<br>(58.6–77.7) | 66.7<br>(48.8–80.8) | 92.1<br>(79.2–97.3) | 70.9<br>(64.8–76.3) |
| Cohen's kappa | 0.031 | 0.500 | 0.319 | 0.242 | 0.803 | 0.357 |
| p value (Fisher's<br>exact) | 1.000 | <0.01 | <0.01 | 0.184 | <0.001 | <0.001 |
| LR+ (95% CI) | 1.08<br>(0.75–1.62) | 3.14<br>(2.20–4.71) | 2.43<br>(1.70–3.65) | 2.00<br>(1.40–3.00) | 22.09<br>(15.46–33.14) | 2.48<br>(1.98–2.97) |
| LR– (95% CI) | 0.93<br>(0.47–1.40) | 0.29<br>(0.14–0.43) | 0.60<br>(0.30–0.90) | 0.50<br>(0.25–0.75) | 0.19<br>(0.09–0.28) | 0.50<br>(0.35–0.65) |
| Youden's index | 0.036 | 0.536 | 0.313 | 0.333 | 0.781 | 0.373 |
| Avg time to<br>assessment | 6m 30s | 11m 48s | 9m 05s | 14m 24s | 13m 30s | 11m 32s |
| Number needed<br>to diagnose | 28.0 | 1.9 | 3.2 | 3.0 | 1.3 | 2.7 |

PPV=positive predictive value; NPV=negative predictive value; LR+=positive likelihood ratio; LR–=negative likelihood ratio; CI=confidence interval. \*Wilson score 95% confidence intervals. Sequential deployment order: Imala Odo (1st) → Ibaro (2nd) → Apojola (3rd) → Abule Titun (4th) → Abule Sikuru (5th). Cochran-Armitage trend tests: accuracy $Z=3.08$ , $p=0.002$ ; specificity $Z=3.00$ , $p=0.003$ ; sensitivity $Z=1.04$ , $p=0.301$ .

**Supplementary Table 4.** Foldscope performance relative to reference standard by laboratory technicians (n=412). Cascade-trained laboratory scientists re-examined 412 intact formalin-preserved glass slides using Foldscope microscopy with smartphone attachment at 340× magnification, blinded to gold-standard results.

|  | Reference standard<br>(+) | Reference standard<br>(-) | Total |
| --- | --- | --- | --- |
| Foldscope (+) | 101 | 2 | 103 |
| Foldscope (-) | 10 | 299 | 309 |
| Total | 111 | 301 | 412 |

**Supplementary Table 5.** Complete Foldscope diagnostic performance metrics — laboratory technicians (n=412).

| Metric | Value | 95% CI |
| --- | --- | --- |
| Sensitivity | 90.99% | 84.21–95.03 |
| Specificity | 99.34% | 97.61–99.88 |
| Positive Predictive Value | 98.06% | 93.19–99.65 |
| Negative Predictive Value | 96.76% | 94.15–98.23 |
| Accuracy | 97.10% | 95.0–98.4 |
| Cohen's kappa | 0.918 | 0.87–0.96 |
| p value (Fisher's exact) | <0.0001 | — |
| Positive likelihood ratio | 136.9 | — |
| Negative likelihood ratio | 0.091 | — |
| Youden's index | 0.90 | — |
| Average time to assessment | 6m 26s | — |
| Number needed to diagnose | 1.1 | — |

*Wilson score method used for all confidence intervals. PPV=positive predictive value; NPV=negative predictive value.*

**Supplementary Table 6.**
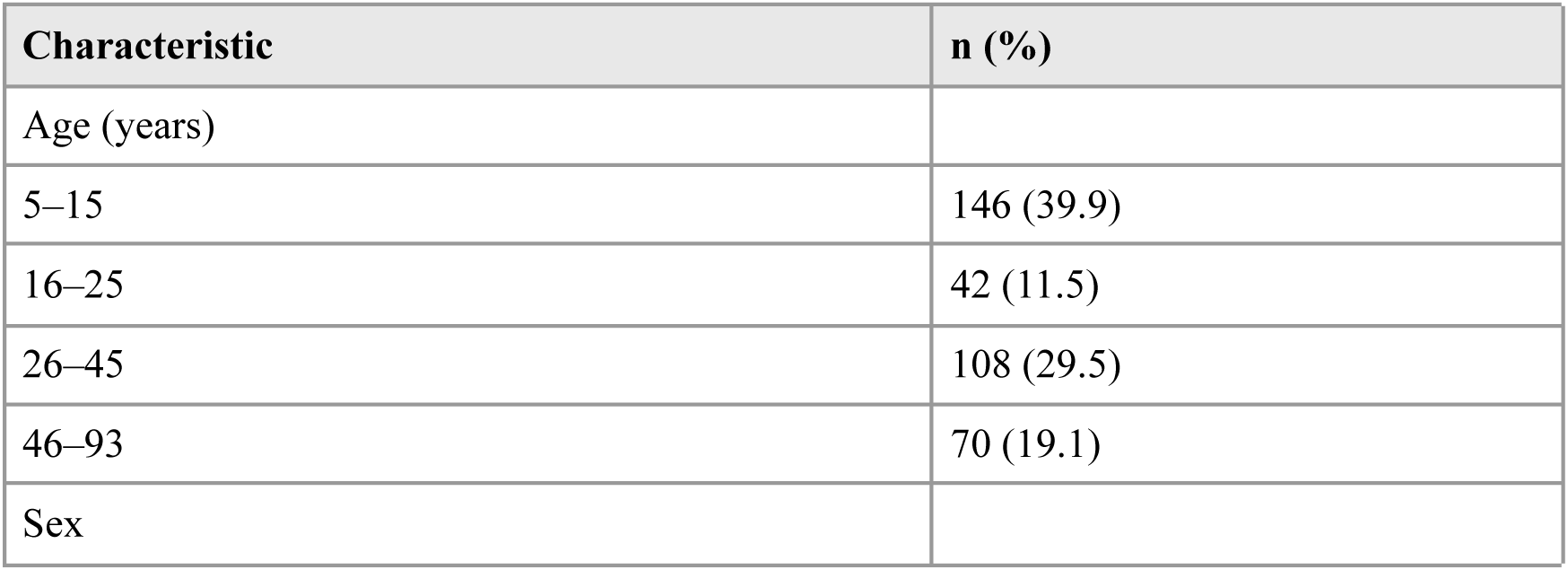
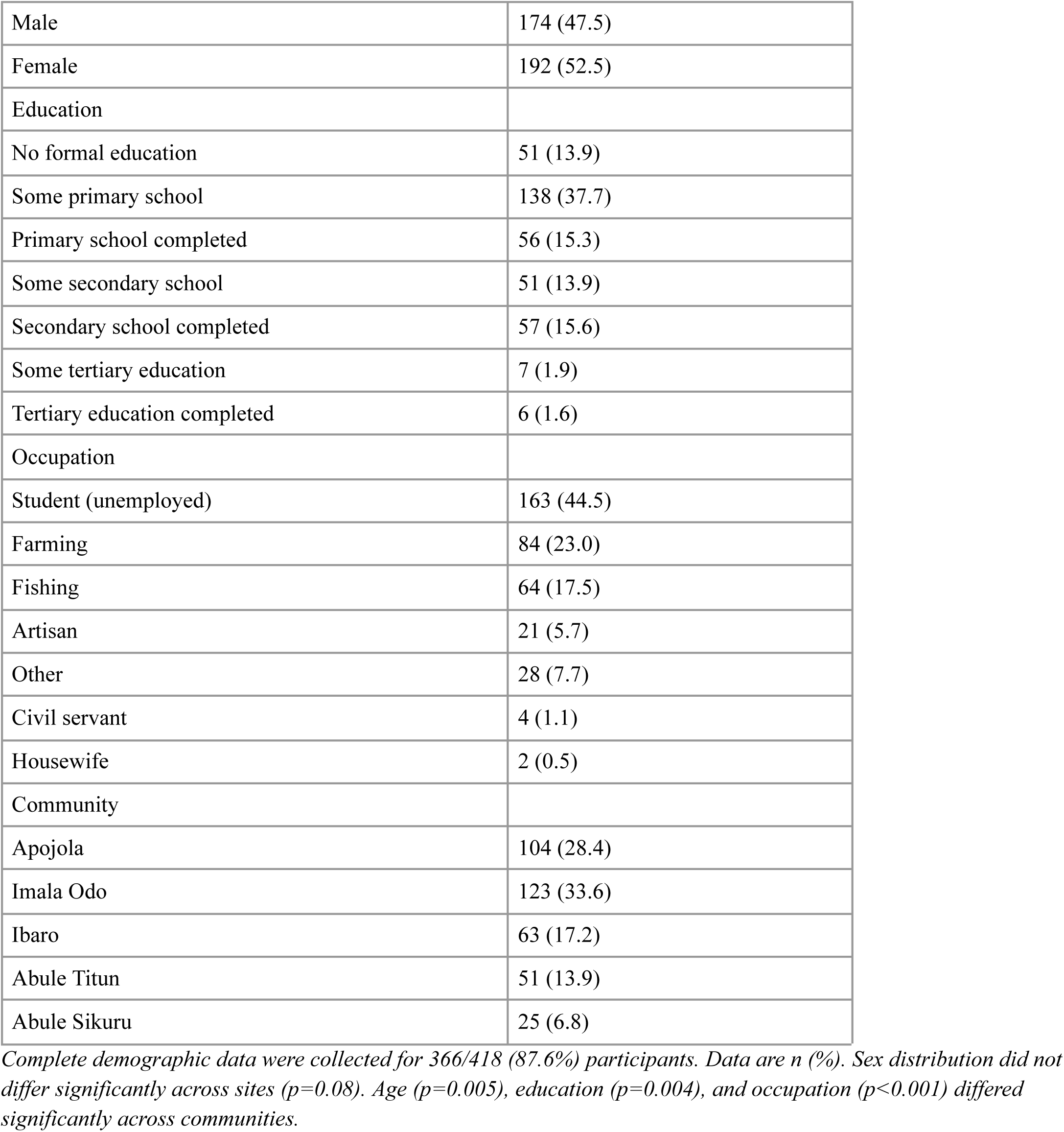
Full demographic characteristics of study participants (n=366).

**Supplementary Table 7.** COREQ 32-item checklist for qualitative studies.

| No. | Item | Guide question/description | Response |
| --- | --- | --- | --- |
| 1 | Interviewer/facilitator | Which author/s conducted the interview? | Dr. Tope Olubodun, Dr. Adeniyi Adebayo, Dr. Adedayo Oludolamu |
| 2 | Credentials | Researcher credentials? | Medical Doctors (MD) |
| 3 | Occupation | Occupation at time of study? | Doctors, Dept. of Community Medicine and Primary Care, Federal Medical Centre, Abeokuta, Ogun State, Nigeria |
| 4 | Gender | Researcher gender? | 2 female, 1 male |
| 5 | Experience and training | Experience or training? | Medical, public health, research; prior work with CHEWs |
| 6 | Relationship established | Relationship established prior to study? | Yes |
| 7 | Participant knowledge of interviewer | What did participants know about the researcher? | Researchers and doctors part of the study; nothing else |
| 8 | Interviewer characteristics | Characteristics reported about interviewer? | Involvement in study proceedings |
| 9 | Methodological orientation | Methodological orientation underpinning the study? | Framework analysis using CFIR with deductive coding (Innovation, Outer Setting, Inner Setting, Individuals) combined with inductive thematic analysis |
| 10 | Sampling | How were participants selected? | Convenience with target sample size |
| 11 | Method of approach | How were participants approached? | Face-to-face |
| 12 | Sample size | How many participants? | 8 total |
| 13 | Non-participation | Refusals or dropouts? | None |
| 14 | Setting of data collection | Where was data collected? | Clinic |
| 15 | Presence of non-participants | Anyone else present? | No |
| 16 | Description of sample | Important sample characteristics? | CHEWs and key informants involved in Ministry of Health NTD control efforts |
| 17 | Interview guide | Were guides provided and pilot tested? | Yes, No |
| 18 | Repeat interviews | Were repeat interviews carried out? | No |
| 19 | Audio/visual recording | Recording used? | Audio recording via SurveyCTO |
| 20 | Field notes | Were field notes made? | Yes |
| 21 | Duration | Duration of interviews? | 1–2 hours |
| 22 | Data saturation | Was data saturation discussed? | Yes |
| 23 | Transcripts returned | Transcripts returned to participants? | Not sent; available upon request |
| 24 | Number of data coders | How many coders? | 3 |
| 25 | Description of coding tree | Coding tree described? | Yes |
| 26 | Derivation of themes | Themes identified in advance or derived from data? | Derived from the data |
| 27 | Software | Software used? | Dedoose |
| 28 | Participant checking | Did participants provide feedback on findings? | Yes |
| 29 | Quotations presented | Participant quotations presented? | Yes; identified by participant number |
| 30 | Data and findings consistent | Consistency between data and findings? | Yes |
| 31 | Clarity of major themes | Major themes clearly presented? | Yes |
| 32 | Clarity of minor themes | Description of diverse cases or minor themes? | Yes |

**Supplementary Table 8.**
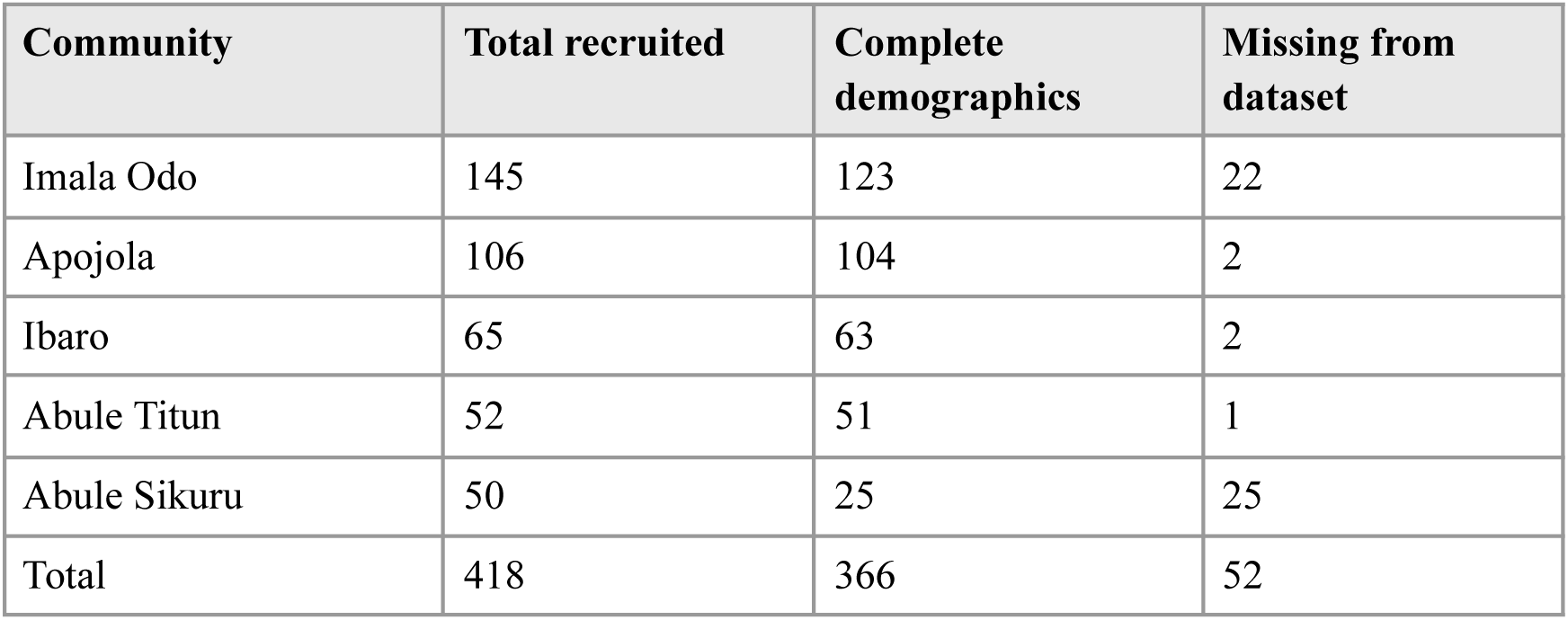
Survey completion and demographic data counts by community.

| Community | Total recruited | Complete demographics | Missing from dataset |
| --- | --- | --- | --- |
| Imala Odo | 145 | 123 | 22 |
| Apojola | 106 | 104 | 2 |
| Ibaro | 65 | 63 | 2 |
| Abule Titun | 52 | 51 | 1 |
| Abule Sikuru | 50 | 25 | 25 |
| Total | 418 | 366 | 52 |

## STARD Checklist | Standards for Reporting Diagnostic Accuracy Studies

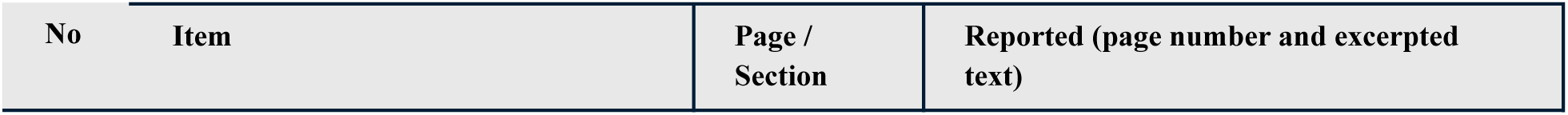
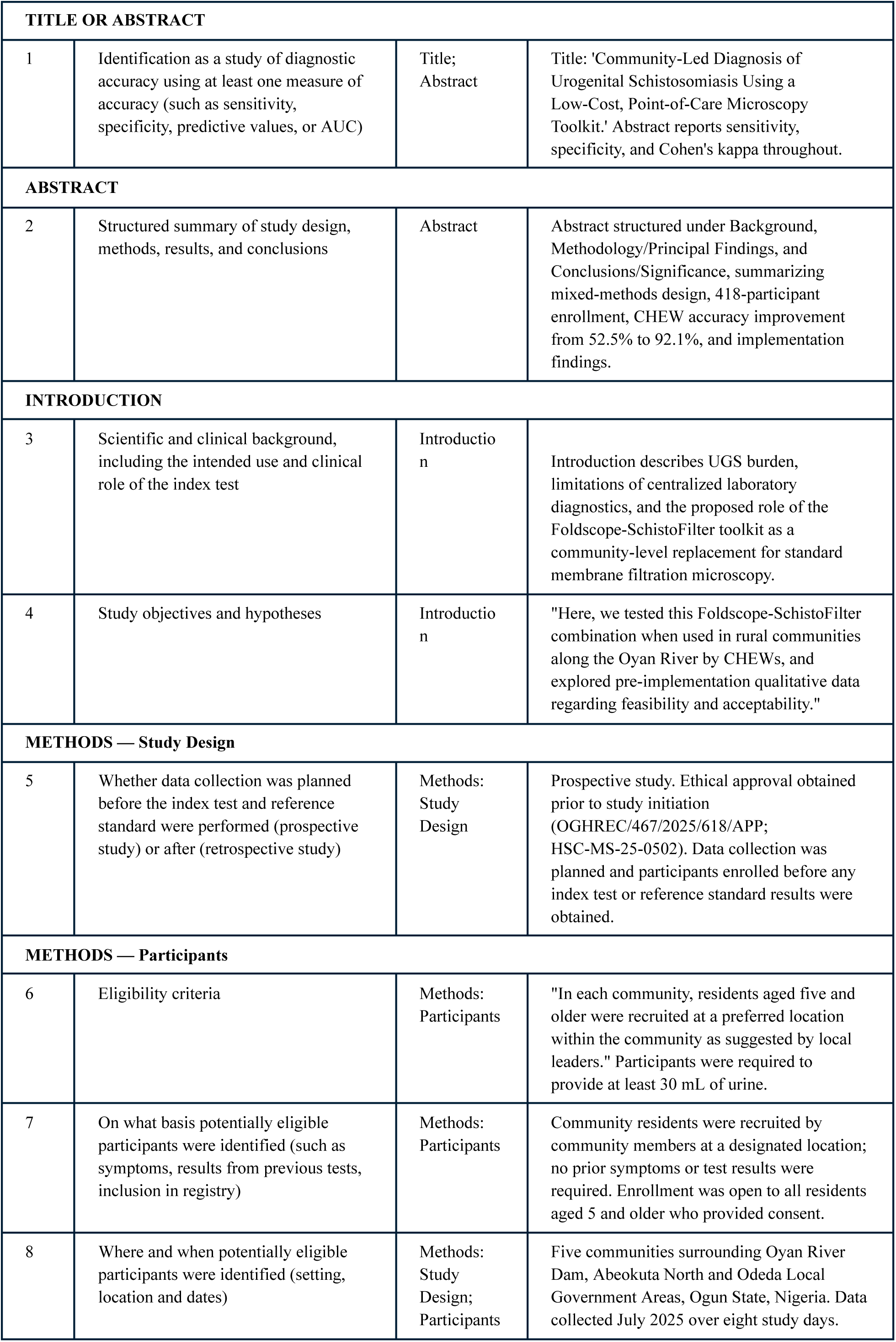
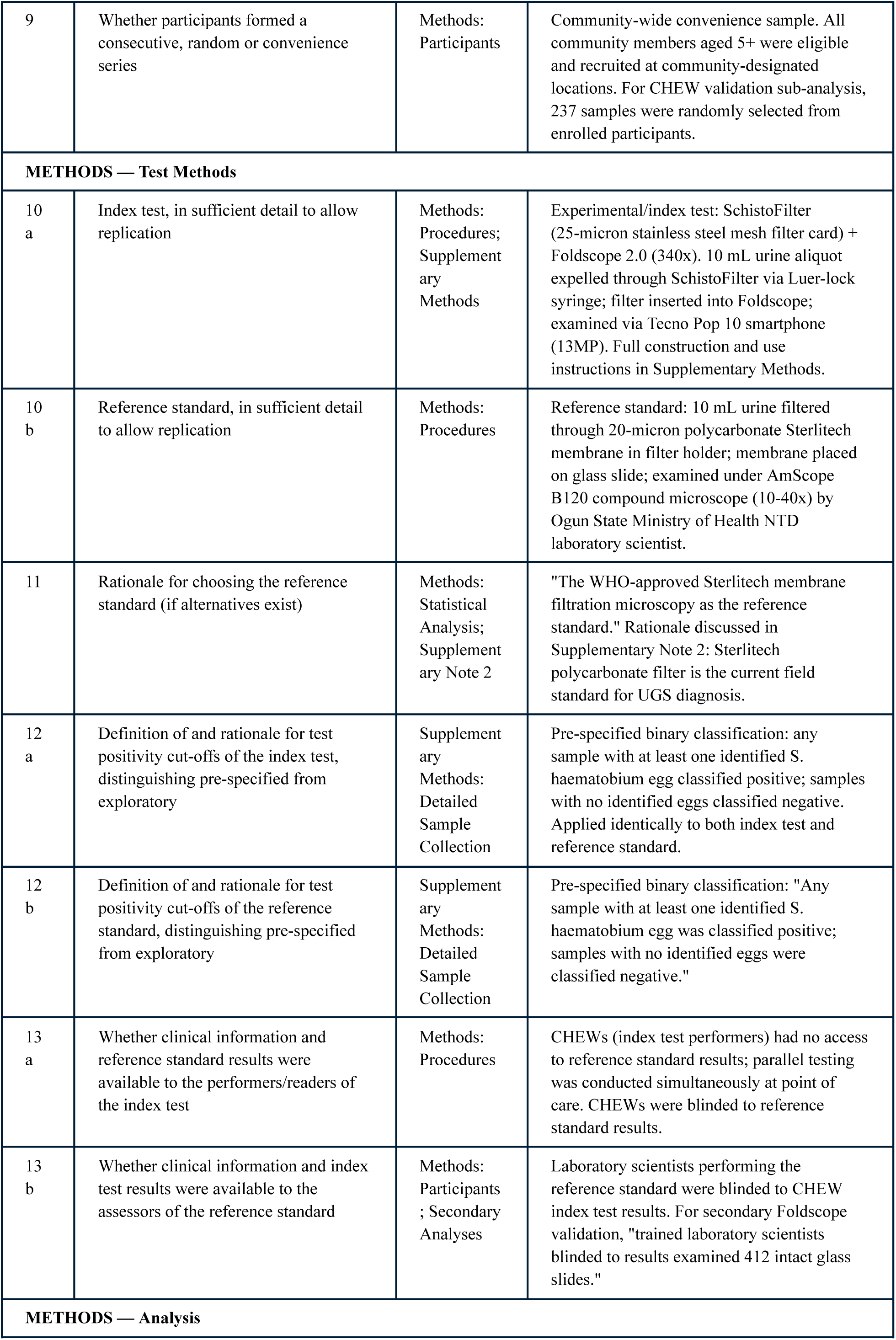
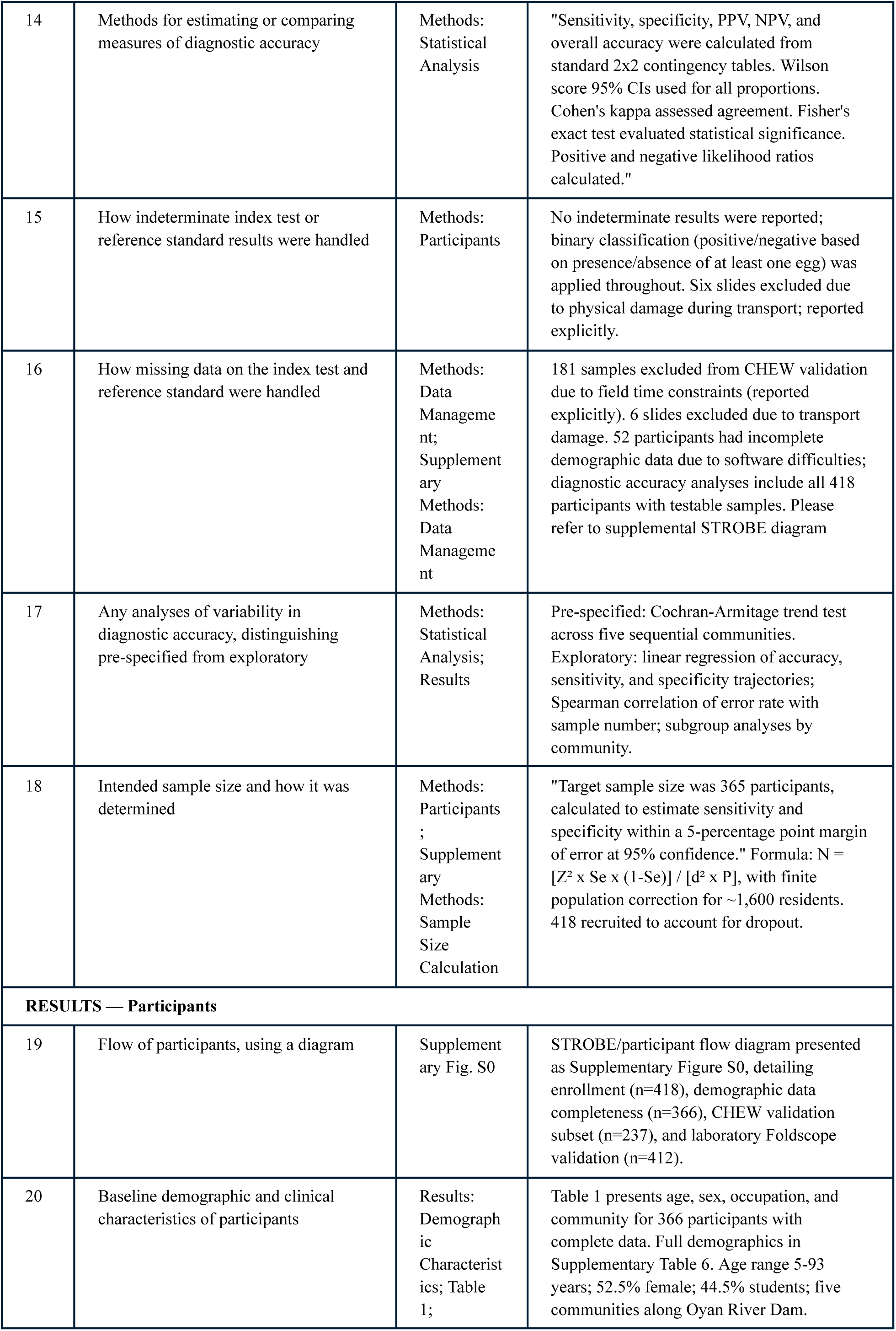
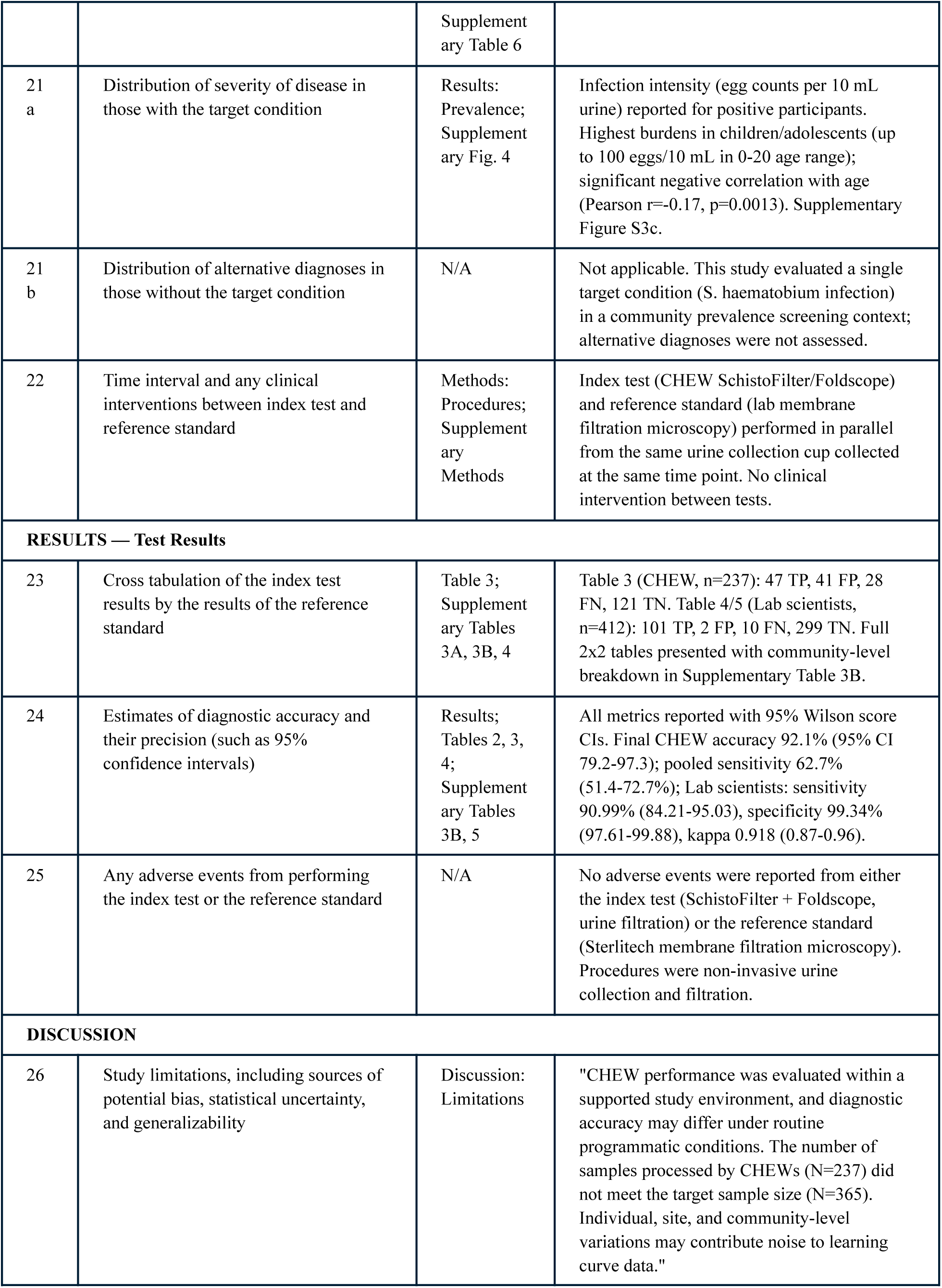
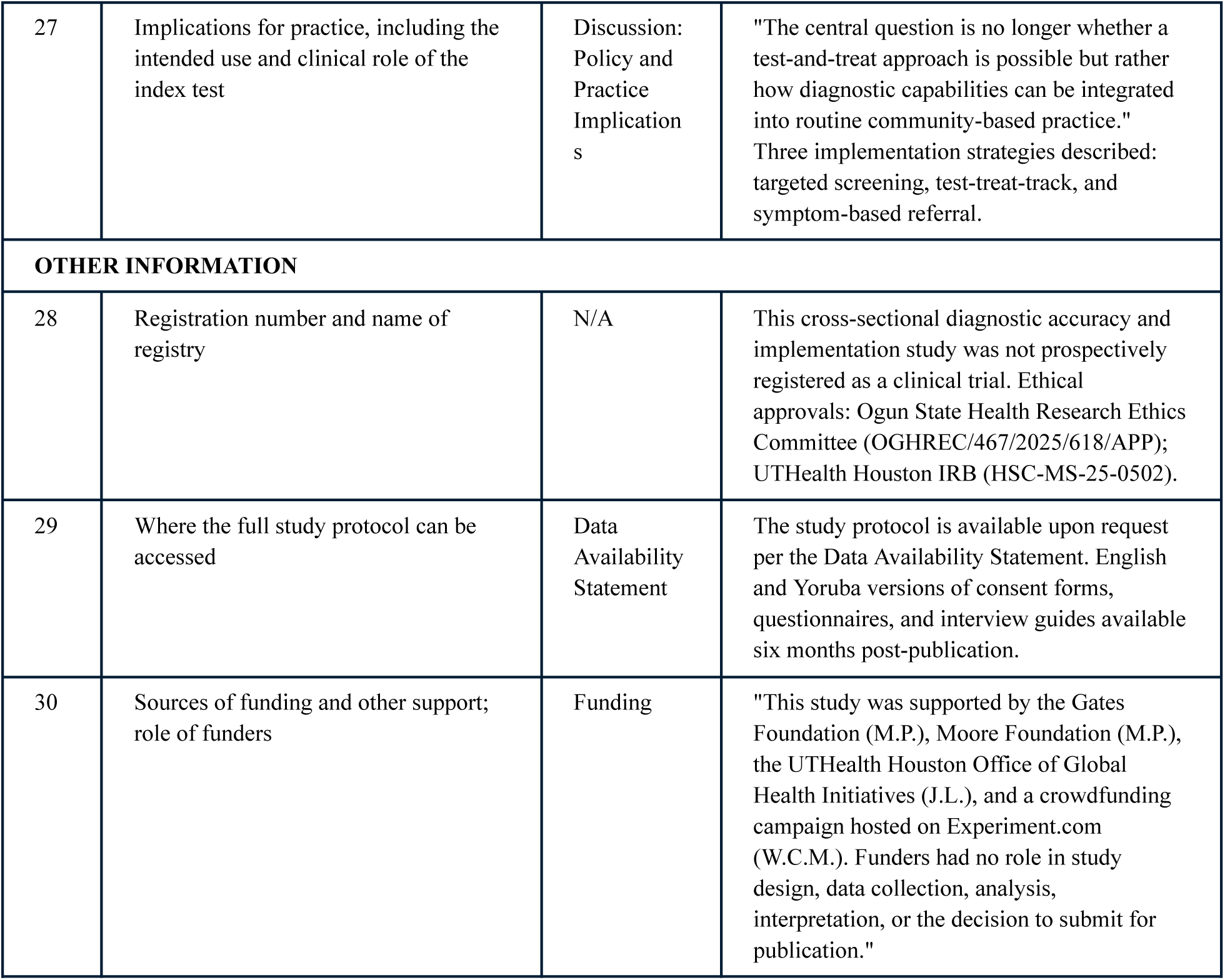

| No | Item | Page / Section | Reported (page number and excerpted text) |
| --- | --- | --- | --- |

| TITLE OR ABSTRACT |  |  |  |
| --- | --- | --- | --- |
| 1 | Identification as a study of diagnostic accuracy using at least one measure of accuracy (such as sensitivity, specificity, predictive values, or AUC) | Title;<br>Abstract | Title: 'Community-Led Diagnosis of Urogenital Schistosomiasis Using a Low-Cost, Point-of-Care Microscopy Toolkit.' Abstract reports sensitivity, specificity, and Cohen's kappa throughout. |
| ABSTRACT |  |  |  |
| 2 | Structured summary of study design, methods, results, and conclusions | Abstract | Abstract structured under Background, Methodology/Principal Findings, and Conclusions/Significance, summarizing mixed-methods design, 418-participant enrollment, CHEW accuracy improvement from 52.5% to 92.1%, and implementation findings. |
| INTRODUCTION |  |  |  |
| 3 | Scientific and clinical background, including the intended use and clinical role of the index test | Introduction | Introduction describes UGS burden, limitations of centralized laboratory diagnostics, and the proposed role of the Foldscope-SchistoFilter toolkit as a community-level replacement for standard membrane filtration microscopy. |
| 4 | Study objectives and hypotheses | Introduction | "Here, we tested this Foldscope-SchistoFilter combination when used in rural communities along the Oyan River by CHEWs, and explored pre-implementation qualitative data regarding feasibility and acceptability." |
| METHODS — Study Design |  |  |  |
| 5 | Whether data collection was planned before the index test and reference standard were performed (prospective study) or after (retrospective study) | Methods:<br>Study Design | Prospective study. Ethical approval obtained prior to study initiation (OGHREC/467/2025/618/APP; HSC-MS-25-0502). Data collection was planned and participants enrolled before any index test or reference standard results were obtained. |
| METHODS — Participants |  |  |  |
| 6 | Eligibility criteria | Methods:<br>Participants | "In each community, residents aged five and older were recruited at a preferred location within the community as suggested by local leaders." Participants were required to provide at least 30 mL of urine. |
| 7 | On what basis potentially eligible participants were identified (such as symptoms, results from previous tests, inclusion in registry) | Methods:<br>Participants | Community residents were recruited by community members at a designated location; no prior symptoms or test results were required. Enrollment was open to all residents aged 5 and older who provided consent. |
| 8 | Where and when potentially eligible participants were identified (setting, location and dates) | Methods:<br>Study Design;<br>Participants | Five communities surrounding Oyan River Dam, Abeokuta North and Odeda Local Government Areas, Ogun State, Nigeria. Data collected July 2025 over eight study days. |
| 9 | Whether participants formed a consecutive, random or convenience series | Methods:<br>Participants | Community-wide convenience sample. All community members aged 5+ were eligible and recruited at community-designated locations. For CHEW validation sub-analysis, 237 samples were randomly selected from enrolled participants. |
| **METHODS — Test Methods** |  |  |  |
| 10<br>a | Index test, in sufficient detail to allow replication | Methods:<br>Procedures;<br>Supplementary<br>Methods | Experimental/index test: SchistoFilter (25-micron stainless steel mesh filter card) + Foldscope 2.0 (340x). 10 mL urine aliquot expelled through SchistoFilter via Luer-lock syringe; filter inserted into Foldscope; examined via Tecno Pop 10 smartphone (13MP). Full construction and use instructions in Supplementary Methods. |
| 10<br>b | Reference standard, in sufficient detail to allow replication | Methods:<br>Procedures | Reference standard: 10 mL urine filtered through 20-micron polycarbonate Sterlitech membrane in filter holder; membrane placed on glass slide; examined under AmScope B120 compound microscope (10-40x) by Ogun State Ministry of Health NTD laboratory scientist. |
| 11 | Rationale for choosing the reference standard (if alternatives exist) | Methods:<br>Statistical Analysis;<br>Supplementary Note 2 | "The WHO-approved Sterlitech membrane filtration microscopy as the reference standard." Rationale discussed in Supplementary Note 2: Sterlitech polycarbonate filter is the current field standard for UGS diagnosis. |
| 12<br>a | Definition of and rationale for test positivity cut-offs of the index test, distinguishing pre-specified from exploratory | Supplementary<br>Methods:<br>Detailed Sample Collection | Pre-specified binary classification: any sample with at least one identified S. haematobium egg classified positive; samples with no identified eggs classified negative. Applied identically to both index test and reference standard. |
| 12<br>b | Definition of and rationale for test positivity cut-offs of the reference standard, distinguishing pre-specified from exploratory | Supplementary<br>Methods:<br>Detailed Sample Collection | Pre-specified binary classification: "Any sample with at least one identified S. haematobium egg was classified positive; samples with no identified eggs were classified negative." |
| 13<br>a | Whether clinical information and reference standard results were available to the performers/readers of the index test | Methods:<br>Procedures | CHEWs (index test performers) had no access to reference standard results; parallel testing was conducted simultaneously at point of care. CHEWs were blinded to reference standard results. |
| 13<br>b | Whether clinical information and index test results were available to the assessors of the reference standard | Methods:<br>Participants<br>; Secondary Analyses | Laboratory scientists performing the reference standard were blinded to CHEW index test results. For secondary Foldscope validation, "trained laboratory scientists blinded to results examined 412 intact glass slides." |
| **METHODS — Analysis** |  |  |  |
| 14 | Methods for estimating or comparing measures of diagnostic accuracy | Methods:<br>Statistical Analysis | "Sensitivity, specificity, PPV, NPV, and overall accuracy were calculated from standard 2x2 contingency tables. Wilson score 95% CIs used for all proportions. Cohen's kappa assessed agreement. Fisher's exact test evaluated statistical significance. Positive and negative likelihood ratios calculated." |
| 15 | How indeterminate index test or reference standard results were handled | Methods:<br>Participants | No indeterminate results were reported; binary classification (positive/negative based on presence/absence of at least one egg) was applied throughout. Six slides excluded due to physical damage during transport; reported explicitly. |
| 16 | How missing data on the index test and reference standard were handled | Methods:<br>Data Management;<br>Supplementary Methods:<br>Data Management | 181 samples excluded from CHEW validation due to field time constraints (reported explicitly). 6 slides excluded due to transport damage. 52 participants had incomplete demographic data due to software difficulties; diagnostic accuracy analyses include all 418 participants with testable samples. Please refer to supplemental STROBE diagram |
| 17 | Any analyses of variability in diagnostic accuracy, distinguishing pre-specified from exploratory | Methods:<br>Statistical Analysis;<br>Results | Pre-specified: Cochran-Armitage trend test across five sequential communities. Exploratory: linear regression of accuracy, sensitivity, and specificity trajectories; Spearman correlation of error rate with sample number; subgroup analyses by community. |
| 18 | Intended sample size and how it was determined | Methods:<br>Participants ;<br>Supplementary Methods:<br>Sample Size Calculation | "Target sample size was 365 participants, calculated to estimate sensitivity and specificity within a 5-percentage point margin of error at 95% confidence." Formula: $N = [Z^2 \times Se \times (1-Se)] / [d^2 \times P]$, with finite population correction for ~1,600 residents. 418 recruited to account for dropout. |
| **RESULTS — Participants** |  |  |  |
| 19 | Flow of participants, using a diagram | Supplementary Fig. S0 | STROBE/participant flow diagram presented as Supplementary Figure S0, detailing enrollment (n=418), demographic data completeness (n=366), CHEW validation subset (n=237), and laboratory Foldscope validation (n=412). |
| 20 | Baseline demographic and clinical characteristics of participants | Results:<br>Demographic Characteristics; Table 1; | Table 1 presents age, sex, occupation, and community for 366 participants with complete data. Full demographics in Supplementary Table 6. Age range 5-93 years; 52.5% female; 44.5% students; five communities along Oyan River Dam. |
|  |  | Supplementary Table 6 |  |
| 21 a | Distribution of severity of disease in those with the target condition | Results: Prevalence; Supplementary Fig. 4 | Infection intensity (egg counts per 10 mL urine) reported for positive participants. Highest burdens in children/adolescents (up to 100 eggs/10 mL in 0-20 age range); significant negative correlation with age (Pearson $r=-0.17$, $p=0.0013$). Supplementary Figure S3c. |
| 21 b | Distribution of alternative diagnoses in those without the target condition | N/A | Not applicable. This study evaluated a single target condition (*S. haematobium* infection) in a community prevalence screening context; alternative diagnoses were not assessed. |
| 22 | Time interval and any clinical interventions between index test and reference standard | Methods: Procedures; Supplementary Methods | Index test (CHEW SchistoFilter/Foldscope) and reference standard (lab membrane filtration microscopy) performed in parallel from the same urine collection cup collected at the same time point. No clinical intervention between tests. |
| **RESULTS — Test Results** |  |  |  |
| 23 | Cross tabulation of the index test results by the results of the reference standard | Table 3; Supplementary Tables 3A, 3B, 4 | Table 3 (CHEW, $n=237$): 47 TP, 41 FP, 28 FN, 121 TN. Table 4/5 (Lab scientists, $n=412$): 101 TP, 2 FP, 10 FN, 299 TN. Full 2x2 tables presented with community-level breakdown in Supplementary Table 3B. |
| 24 | Estimates of diagnostic accuracy and their precision (such as 95% confidence intervals) | Results; Tables 2, 3, 4; Supplementary Tables 3B, 5 | All metrics reported with 95% Wilson score CIs. Final CHEW accuracy 92.1% (95% CI 79.2-97.3); pooled sensitivity 62.7% (51.4-72.7%); Lab scientists: sensitivity 90.99% (84.21-95.03), specificity 99.34% (97.61-99.88), kappa 0.918 (0.87-0.96). |
| 25 | Any adverse events from performing the index test or the reference standard | N/A | No adverse events were reported from either the index test (SchistoFilter + Foldscope, urine filtration) or the reference standard (Sterlitech membrane filtration microscopy). Procedures were non-invasive urine collection and filtration. |
| **DISCUSSION** |  |  |  |
| 26 | Study limitations, including sources of potential bias, statistical uncertainty, and generalizability | Discussion: Limitations | "CHEW performance was evaluated within a supported study environment, and diagnostic accuracy may differ under routine programmatic conditions. The number of samples processed by CHEWs ($N=237$) did not meet the target sample size ($N=365$). Individual, site, and community-level variations may contribute noise to learning curve data." |
| 27 | Implications for practice, including the intended use and clinical role of the index test | Discussion: Policy and Practice Implications | "The central question is no longer whether a test-and-treat approach is possible but rather how diagnostic capabilities can be integrated into routine community-based practice."<br>Three implementation strategies described: targeted screening, test-treat-track, and symptom-based referral. |
| **OTHER INFORMATION** |  |  |  |
| 28 | Registration number and name of registry | N/A | This cross-sectional diagnostic accuracy and implementation study was not prospectively registered as a clinical trial. Ethical approvals: Ogun State Health Research Ethics Committee (OGHREC/467/2025/618/APP); UTHealth Houston IRB (HSC-MS-25-0502). |
| 29 | Where the full study protocol can be accessed | Data Availability Statement | The study protocol is available upon request per the Data Availability Statement. English and Yoruba versions of consent forms, questionnaires, and interview guides available six months post-publication. |
| 30 | Sources of funding and other support; role of funders | Funding | "This study was supported by the Gates Foundation (M.P.), Moore Foundation (M.P.), the UTHealth Houston Office of Global Health Initiatives (J.L.), and a crowdfunding campaign hosted on Experiment.com (W.C.M.). Funders had no role in study design, data collection, analysis, interpretation, or the decision to submit for publication." |

## Supplementary Statistical Appendix

### Simple linear regression: infection intensity vs. age

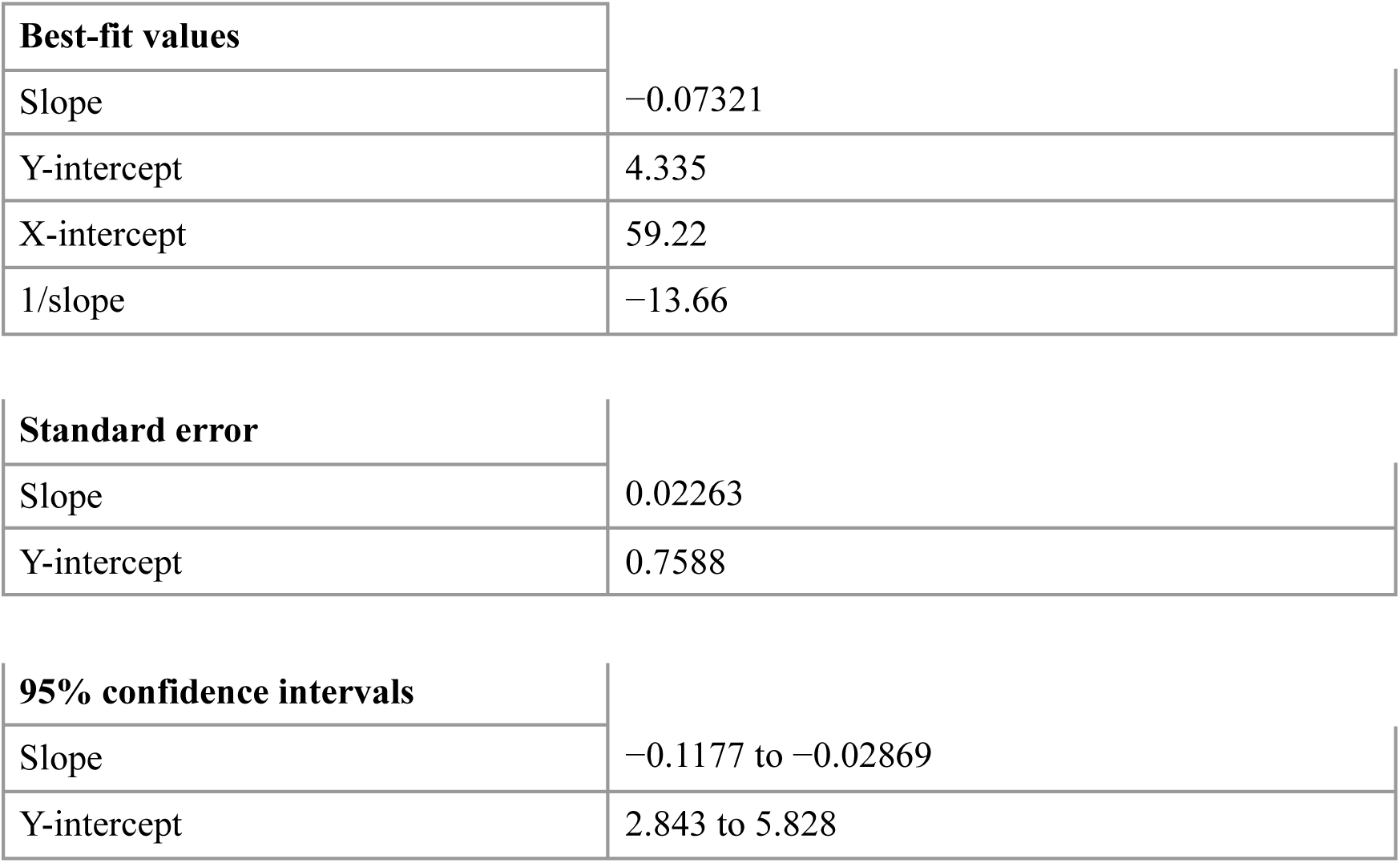
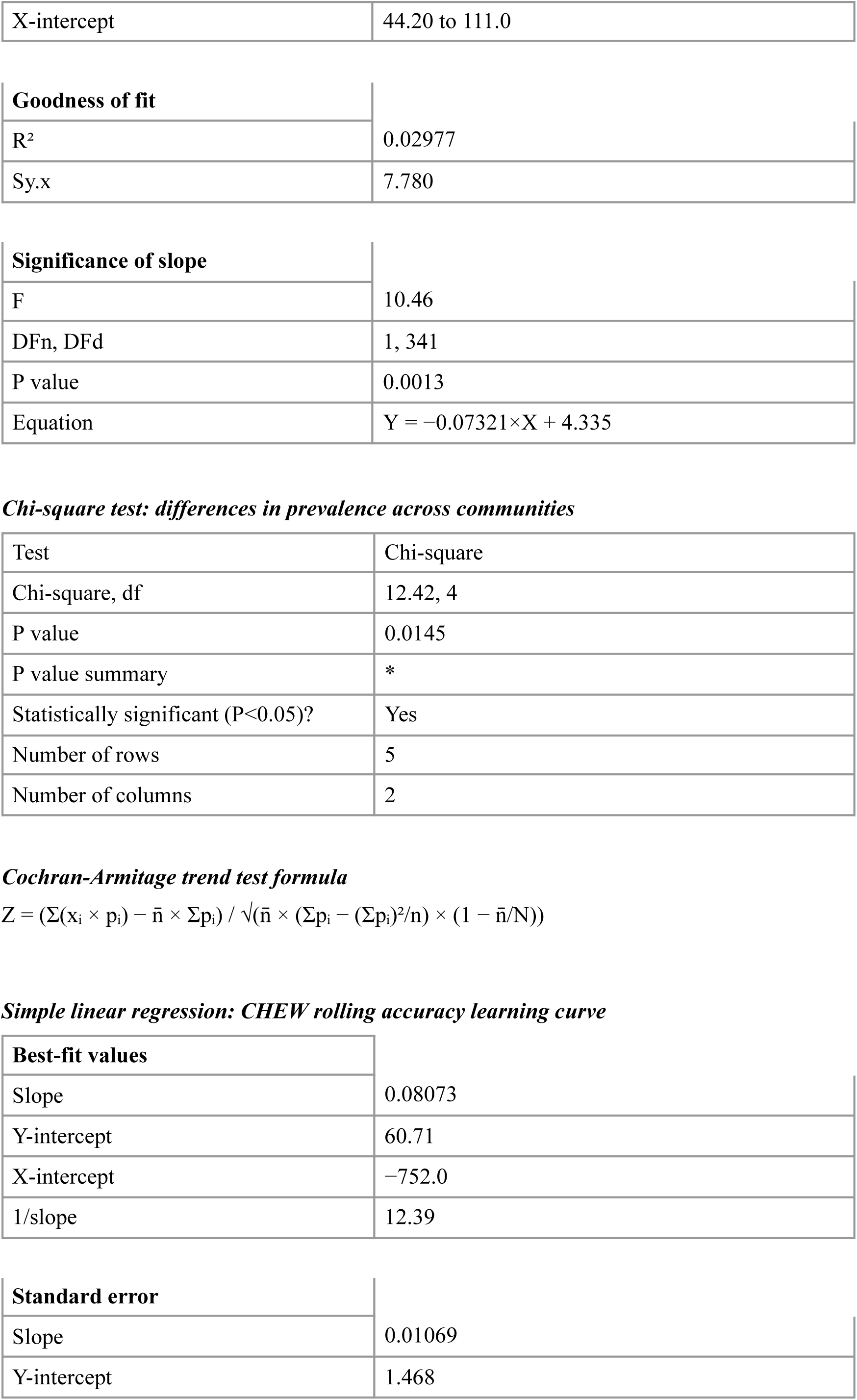
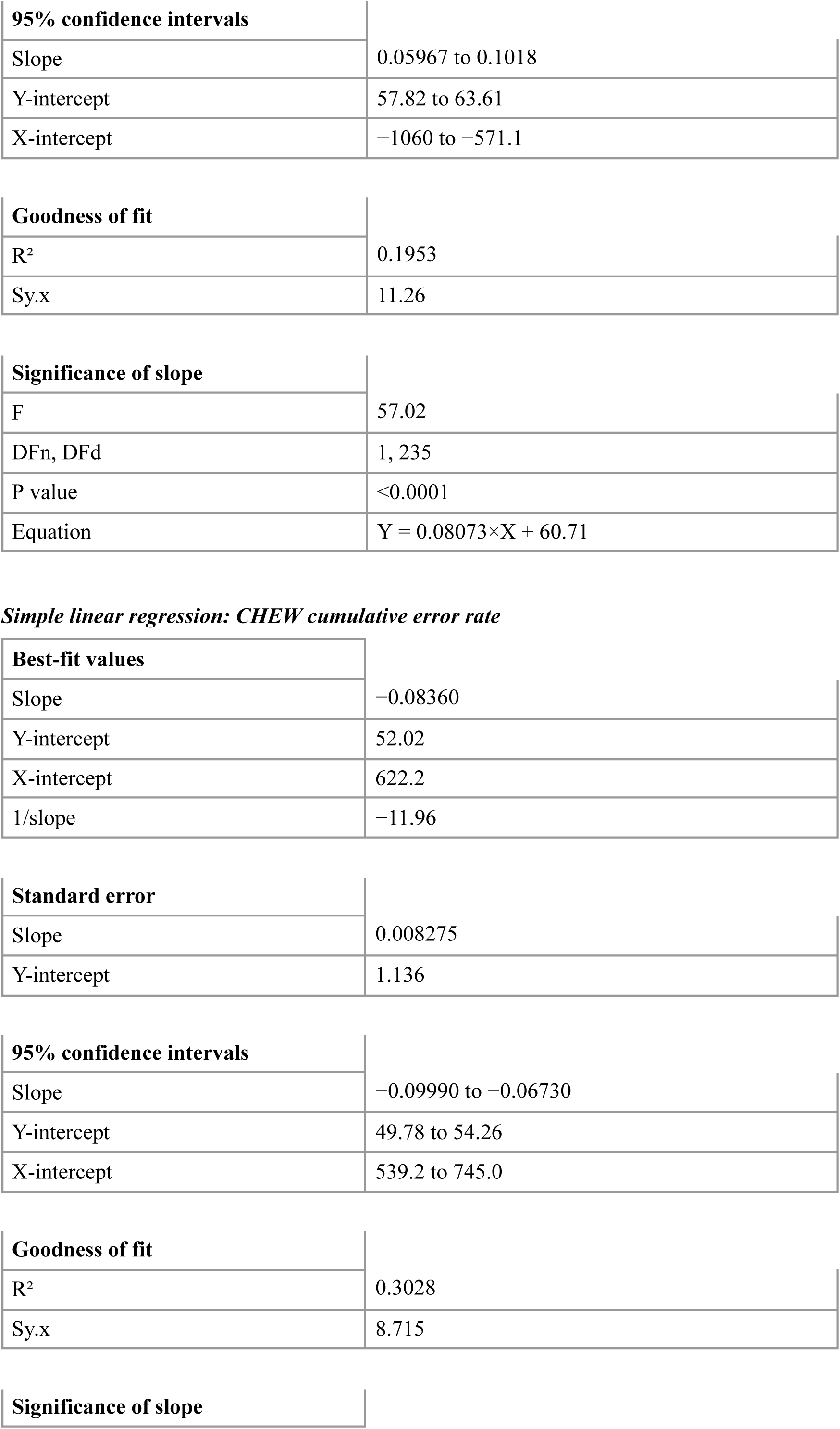
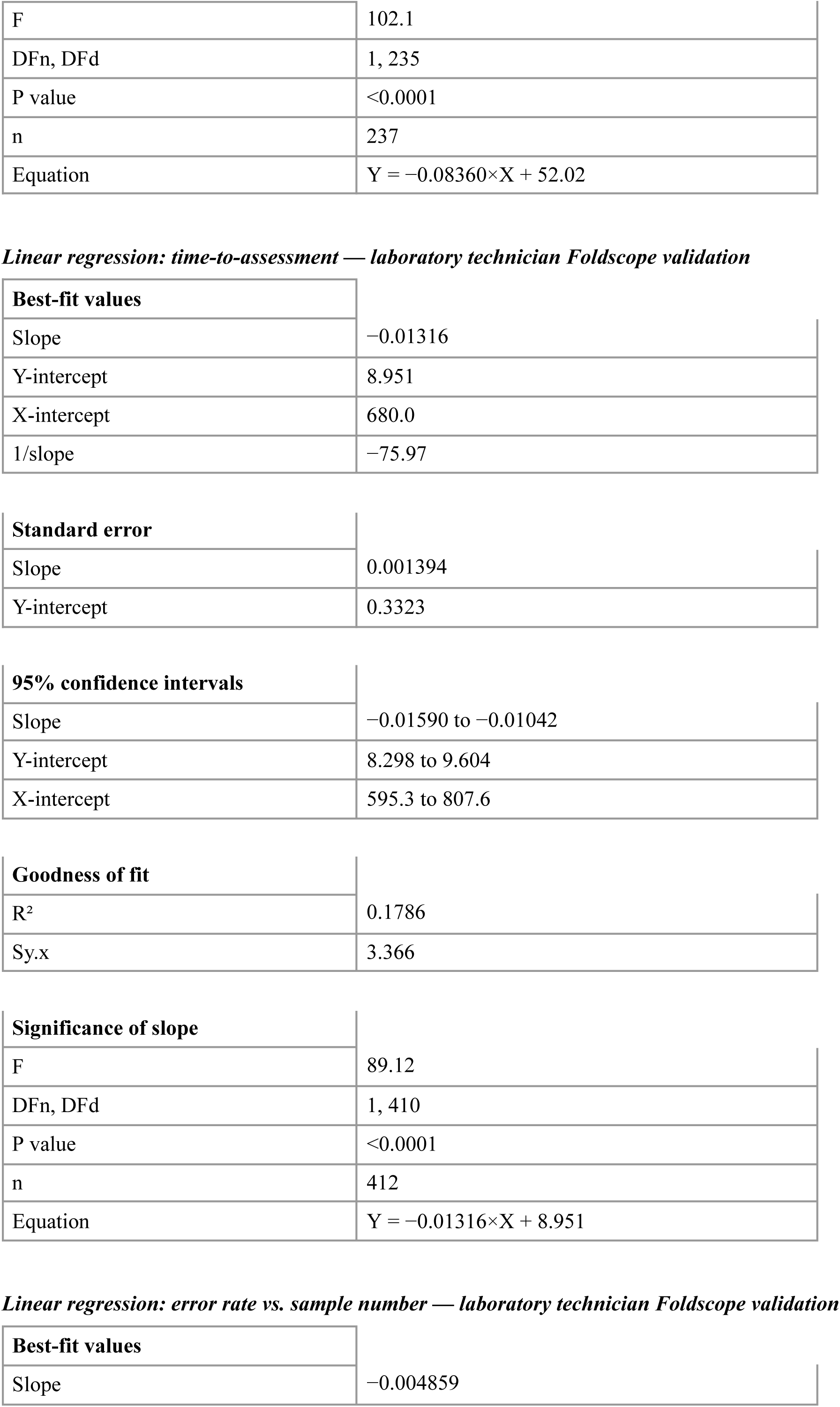
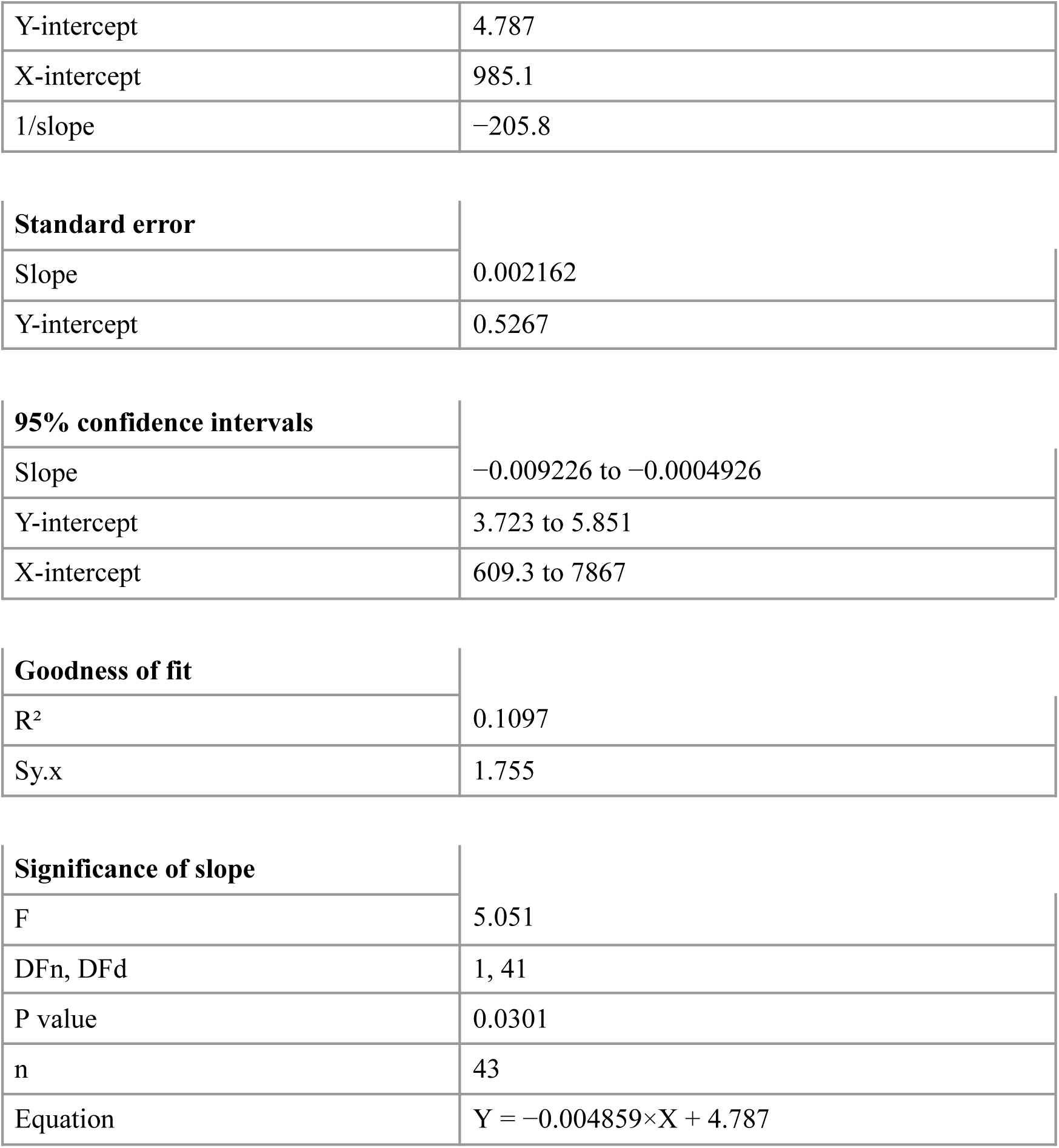

Supplementary Statistical Appendix *Simple linear regression: infection intensity vs. age*
| Best-fit values |  |
| --- | --- |
| Slope | −0.07321 |
| Y-intercept | 4.335 |
| X-intercept | 59.22 |
| 1/slope | −13.66 |

| Standard error |  |
| --- | --- |
| Slope | 0.02263 |
| Y-intercept | 0.7588 |

| 95% confidence intervals |  |
| --- | --- |
| Slope | −0.1177 to −0.02869 |
| Y-intercept | 2.843 to 5.828 |
| X-intercept | 44.20 to 111.0 |
| **Goodness of fit** |  |
| R<sup>2</sup> | 0.02977 |
| Sy.x | 7.780 |
| **Significance of slope** |  |
| F | 10.46 |
| DFn, DFd | 1, 341 |
| P value | 0.0013 |
| Equation | $Y = -0.07321 \times X + 4.335$ |
| ***Chi-square test: differences in prevalence across communities*** |  |
| Test | Chi-square |
| Chi-square, df | 12.42, 4 |
| P value | 0.0145 |
| P value summary | * |
| Statistically significant (P<0.05)? | Yes |
| Number of rows | 5 |
| Number of columns | 2 |
| **Best-fit values** |  |
| Slope | 0.08073 |
| Y-intercept | 60.71 |
| X-intercept | -752.0 |
| 1/slope | 12.39 |
| **Standard error** |  |
| Slope | 0.01069 |
| Y-intercept | 1.468 |
| **95% confidence intervals** |  |
| Slope | 0.05967 to 0.1018 |
| Y-intercept | 57.82 to 63.61 |
| X-intercept | −1060 to −571.1 |
| **Goodness of fit** |  |
| R<sup>2</sup> | 0.1953 |
| Sy.x | 11.26 |
| **Significance of slope** |  |
| F | 57.02 |
| DFn, DFd | 1, 235 |
| P value | <0.0001 |
| Equation | $Y = 0.08073 \times X + 60.71$ |
| ***Simple linear regression: CHEW cumulative error rate*** |  |
| **Best-fit values** |  |
| Slope | −0.08360 |
| Y-intercept | 52.02 |
| X-intercept | 622.2 |
| 1/slope | −11.96 |
| **Standard error** |  |
| Slope | 0.008275 |
| Y-intercept | 1.136 |
| **95% confidence intervals** |  |
| Slope | −0.09990 to −0.06730 |
| Y-intercept | 49.78 to 54.26 |
| X-intercept | 539.2 to 745.0 |
| **Goodness of fit** |  |
| R<sup>2</sup> | 0.3028 |
| Sy.x | 8.715 |
| **Significance of slope** |  |
| F | 102.1 |
| DFn, DFd | 1, 235 |
| P value | <0.0001 |
| n | 237 |
| Equation | $Y = -0.08360 \times X + 52.02$ |
| ***Linear regression: time-to-assessment — laboratory technician Foldscope validation*** |  |
| **Best-fit values** |  |
| Slope | -0.01316 |
| Y-intercept | 8.951 |
| X-intercept | 680.0 |
| 1/slope | -75.97 |
| **Standard error** |  |
| Slope | 0.001394 |
| Y-intercept | 0.3323 |
| **95% confidence intervals** |  |
| Slope | -0.01590 to -0.01042 |
| Y-intercept | 8.298 to 9.604 |
| X-intercept | 595.3 to 807.6 |
| **Goodness of fit** |  |
| R<sup>2</sup> | 0.1786 |
| Sy.x | 3.366 |
| **Significance of slope** |  |
| F | 89.12 |
| DFn, DFd | 1, 410 |
| P value | <0.0001 |
| n | 412 |
| Equation | $Y = -0.01316 \times X + 8.951$ |
| ***Linear regression: error rate vs. sample number — laboratory technician Foldscope validation*** |  |
| **Best-fit values** |  |
| Slope | -0.004859 |
| Y-intercept | 4.787 |
| X-intercept | 985.1 |
| 1/slope | -205.8 |
| **Standard error** |  |
| Slope | 0.002162 |
| Y-intercept | 0.5267 |
| **95% confidence intervals** |  |
| Slope | -0.009226 to -0.0004926 |
| Y-intercept | 3.723 to 5.851 |
| X-intercept | 609.3 to 7867 |
| **Goodness of fit** |  |
| R<sup>2</sup> | 0.1097 |
| Sy.x | 1.755 |
| **Significance of slope** |  |
| F | 5.051 |
| DFn, DFd | 1, 41 |
| P value | 0.0301 |
| n | 43 |
| Equation | $Y = -0.004859 \times X + 4.787$ |

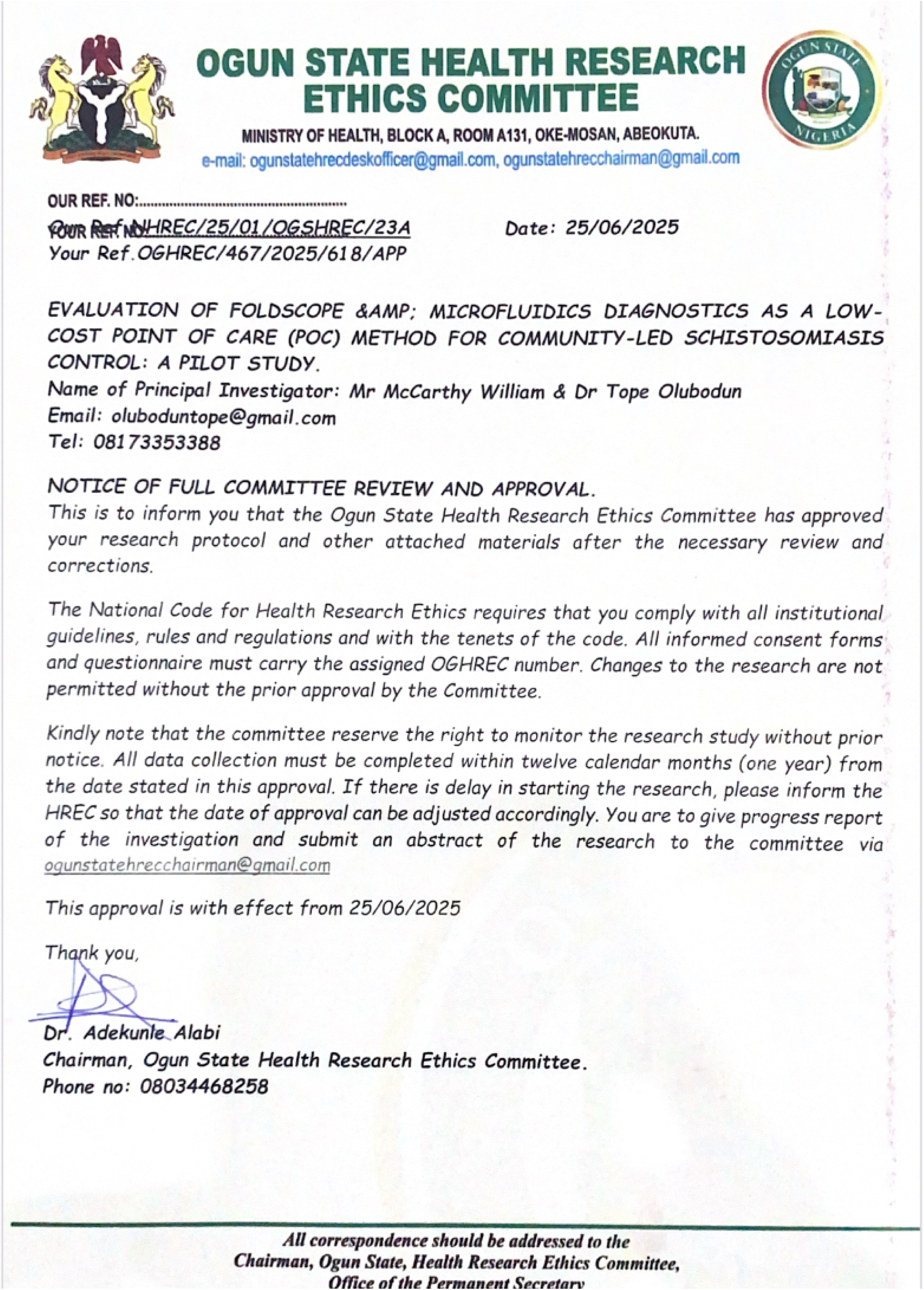

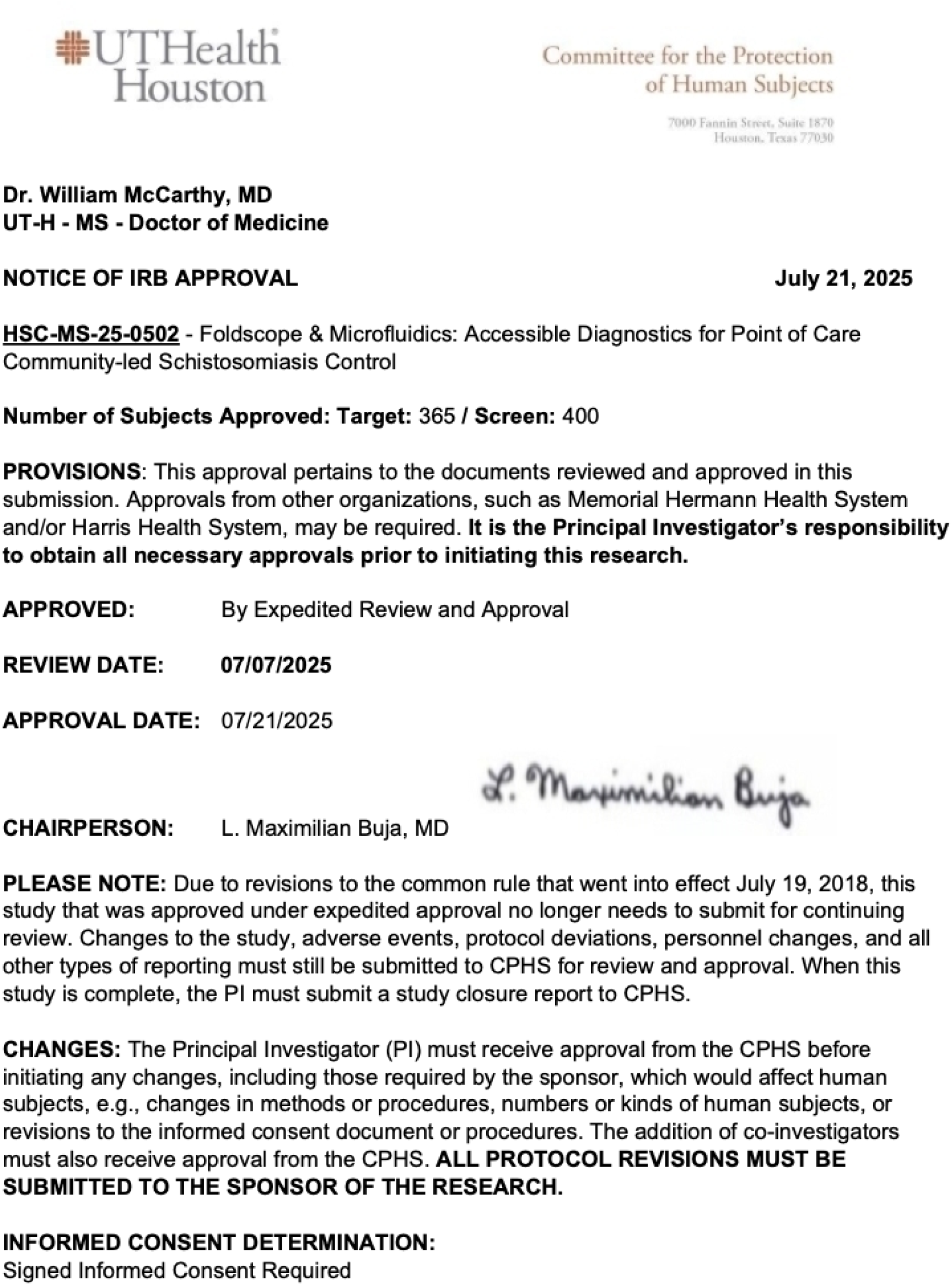

## Notes

### Clinical Protocols

https://dx.doi.org/10.17504/protocols.io.4r3l2dpkjg1y/v1

https://www.protocols.io/private/1914F3331F4911F19C7D0A58A9FEAC02

### Funding Statement

Yes

